# Persistent somatic symptoms are key to individual illness perception at one year after COVID-19

**DOI:** 10.1101/2022.09.05.22279602

**Authors:** Katharina Hüfner, Piotr Tymoszuk, Sabina Sahanic, Anna Luger, Anna Boehm, Alex Pizzini, Christoph Schwabl, Sabine Koppelstätter, Katharina Kurz, Malte Asshoff, Birgit Mosheimer-Feistritzer, Bernhard Pfeifer, Verena Rass, Andrea Schroll, Sarah Iglseder, Alexander Egger, Ewald Wöll, Günter Weiss, Raimund Helbok, Gerlig Widmann, Thomas Sonnweber, Ivan Tancevski, Barbara Sperner-Unterweger, Judith Löffler-Ragg

## Abstract

**Background:** Sequelae of Coronavirus disease 2019 (COVID-19) were investigated by both patient-initiated and academic initiatives. Patient’s subjective illness perceptions might differ from physician’s clinical assessment results. Herein, we explored factors influencing patient’s perception during COVID-19 recovery.

**Methods:** Participants of the prospective observation CovILD study with persistent somatic symptoms or cardiopulmonary findings at the clinical follow-up one year after COVID-19 were analyzed (n = 74). Explanatory variables included baseline demographic and comorbidity data, COVID-19 course and one-year follow-up data of persistent somatic symptoms, physical performance, lung function testing (LFT), chest computed tomography (CT) and trans-thoracic echocardiography (TTE). Factors affecting illness perception (Brief Illness Perception Questionnaire, BIPQ) were identified by penalized multi-parameter regression and unsupervised clustering.

**Results:** In modeling, 47% of overall illness perception variance at one year after COVID-19 was attributed to fatigue intensity, reduced physical performance, hair loss and baseline respiratory comorbidity. Overall illness perception was independent of LFT results, pulmonary lesions in CT or heart abnormality in TTE. As identified by clustering, persistent somatic symptom count, fatigue, diminished physical performance, dyspnea, hair loss and sleep problems at the one-year follow-up and severe acute COVID-19 were associated with the BIPQ domains of concern, emotional representation, complaints, disease timeline and consequences.

**Conclusion:** Persistent somatic symptoms rather than clinical assessment results, revealing lung and heart abnormalities, impact on severity and quality of illness perception at one year after COVID-19 and may foster unhelpful coping mechanisms. Besides COVID-19 severity, individual illness perception should be taken into account when allocating rehabilitation and psychological therapy resources.

**Study registration:** ClinicalTrials.gov: NCT04416100.

## Introduction

A sizable fraction of coronavirus disease 2019 (COVID-19) patients is affected by protracted somatic symptoms, cardiopulmonary pathology and mental health disorders (1–9). Persistent COVID-19-related symptoms have been initially described by patient and social media initiatives (10) and subsequently, recognized as the ‘post COVID-19 condition’ by the clinical community (1,2,6–8). Yet, the patients’ and clinicians’ characteristics were not always consistent, which was observed also in other conditions, such as chronic obstructive pulmonary disease (11,12) or functional disorders (13). For post COVID-19 condition, the matter is further complicated by a broad range of patient-reported manifestations and widespread character of the disease (2,7). Since it is becoming increasingly evident that many COVID-19 convalescents experience prolonged severe individual suffering (5,7,9,14), and pose a significant healthcare and socioeconomic challenge (6), it is critically important to characterize and understand individual illness perception following COVID-19.

Illness perceptions can be divided into cognitive or emotional components (15). The cognitive representations include self-perceived consequences, expected duration, personal control, expected effect of treatment, symptom perception and understanding of the disease. The emotional components encompass patient’s concerns and emotions e.g. fear, anger or distress associated with the disease (15,16). According to the common-sense model of self regulation (CSM), a theoretical framework enabling understanding of how people cope with threats to their health, such illness perceptions are influenced by situational stimuli such as symptoms, health information and patient’s knowledge (17–19). The individual illness perception was shown to influence disease coping, adjustment to adverse life events or chronic conditions and compliance with prevention, treatment and rehabilitation (20,21), also in context of acute COVID-19 (22–24). Finally, severe illness perception including the emotional response, concerns and consequence components was found associated with anxiety, depression and stress both in the general population and in COVID-19 patients (25–28).

Herein, we investigated severity of overall illness perception and the illness perception components in a cross-sectional COVID-19 convalescent collective (1–4) with incomplete somatic symptom resolution or COVID-19-related lung and heart pathology in the follow-up clinical assessment at one year after diagnosis.

## Methods

### Study design and approval

Participants of the longitudinal observation CovILD study (ClinicalTrials.gov: NCT04416100) were recruited between April and June 2020 at three Austrian clinical centers (1,3,4). The study inclusion criteria were age *≥* 18 years and symptomatic PCR-confirmed SARS-CoV-2 infection. The follow-up visits were scheduled at two, three, six months and one year after COVID-19 diagnosis. A subset of participants (n = 74) was analyzed who completed the Brief Illness Perception Questionnaire (BIPQ) (15) and displayed (1) COVID-19-related persistent somatic symptoms or (2) any abnormality in chest computed tomography (CT) or (3) any lung function testing (LFT) deficits or (4) any heart abnormality in trans-thoracic echocardiography (TTE) at the one-year follow-up (**Figure 1**).

**Figure 1.**
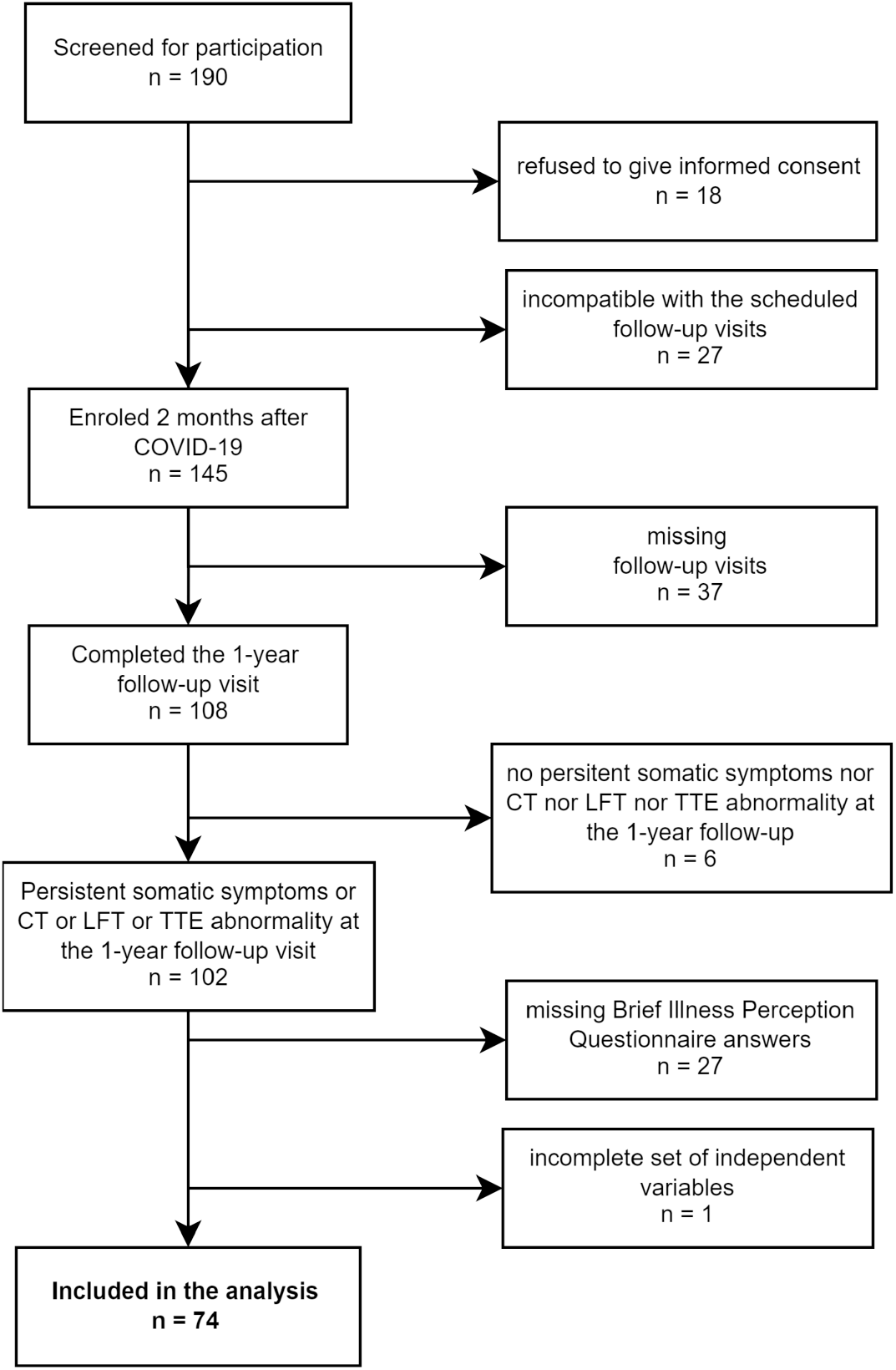
Flow diagram of study enrollment and analysis inclusion process. CT: computed tomography of the chest; LFT: lung function testing, TTE: trans-thoracic echocardiography.

The study was performed in accordance with the Declaration of Helsinki and the European Data Policy. All participants gave written informed consent. The study protocol was approved by the ethics committee at the Medical University of Innsbruck (approval number 1103/2020).

### Procedures

For full descriptions of procedures and variables, see **Supplementary Methods** and **Supplementary Table S1**.

Baseline clinical and acute COVID-19 data were recorded retrospectively at the two-month follow-up based on the patient’s interview and electronic patient records (1). Study participants were classified as ambulatory (outpatient, WHO grade 1 - 2), moderate (hospitalized, without oxygen therapy or mask/nasal prongs oxygen, WHO 3 - 4) and severe COVID-19 survivors (hospitalized with non-invasive ventilation or high-flow oxygen or mechanical ventilation, WHO 5 - 7).

Physical performance was rated with the Eastern Cooperative Oncology Group scale (ECOG). Dyspnea was scored with the Modified Medical British Research Council scale (mMRC). Fatigue at one-year follow-up was rated with likert and bimodal Chalder’s Fatigue Scales (CFS) (29,30). Exertional capacity at the one-year follow-up was assessed by six-minute walking distance (SMWD) test and compared with the reference values (31).

The following COVID-19-related persistent somatic symptoms at the one-year follow-up were analyzed: reduced physical performance (ECOG *≥* 1), dyspnea (mMRC *≥* 1), self-reported cough (yes/no item), self-reported sleep problems (yes/no), self-reported night sweating (yes/no), self-reported hyposmia or anosmia (yes/no), self-reported dermatological symptoms (yes/no), self-reported gastrointestinal symptoms (yes/no), self-reported hair loss (yes/no), significant fatigue (bimodal CFS *≥* 4).

LFT abnormality was defined as at least one parameter < 80% (forced vital capacity [FVC], forced expiratory volume in 1 second [FEV1], total lung capacity, diffusion lung capacity for carbon monoxide) or < 70% (FEV1:FVC ratio) of the reference value (4). CT images were evaluated with the Fleischner Society glossary terms (32) and the CT severity score (1,3,4). Blood biomarkers encompassed hemoglobin and parameters of iron turnover, inflammation and coagulation.

Illness perception was investigated with the 8-item BIPQ (Q1 - Q8) (15). Each item was rated with an 11-point likert scale. The illness perception score was defined as a sum of all BIPQ items, with the negative items Q3, Q4 and Q7 inverted.

## Analysis endpoints

The primary analysis endpoints were illness perception score values and its influencing factors among demographic, clinical and somatic symptom data at the one-year after COVID-19 in convalescents affected by persistent somatic symptoms or residual cardiopulmonary findings. The secondary analysis endpoints were particular BIPQ component scores and their influencing factors.

## Statistical analysis

Details of statistical analysis are provided in **Supplementary Methods**. Data analysis was done with R version 4.2.0 (R Foundation for Statistical Computing). Differences in categorical variable distribution were assessed by *χ*^2^ test with Cramer V effect size statistic. Statistical significance for numerical variables was investigated by Mann-Whitney test with Wilcoxon r effect size statistic or Kruskal-Wallis test with *η*^2^ effect size statistic (33). Correlations were investigated by Spearman’s test.

Multi-parameter modeling was done with the Elastic Net (package *glmnet*) (34,35), LASSO (least absolute shrinkage and selection operator; package *glmnet*) (35,36) and Bayesian LASSO (package *monomvn*) (37,38) algorithms. The response, illness perception score, was square-root transformed to guarantee normality. Both first and second order terms of numeric explanatory variables were included in the models. Numeric explanatory and response variables were Z-score normalized. The optimal *λ* for Elastic Net and LASSO were obtained by 200-repeats 10-fold cross-validation (CV). The ‘sparsity’ parameter in Bayesian LASSO was found by 10-repeats 10-fold cross-validation (package *caret*) (39). Explained variance (*R*^2^) and model root mean squared error (RMSE) were assessed in the entire data set and 10-repeats 10-fold CV (package *caret*) (39). Elastic Net and LASSO coefficients were calculated for the optimal *λ* values. Bayesian LASSO coefficients were calculated as medians over all algorithm iterations (37). Variables with non-zero coefficients in all three models were deemed key factors for the illness perception score.

Clustering by the BIPQ items was accomplished with the PAM algorithm (partitioning around medoids; Euclidean distance) (40,41).

## Results

### Study cohort at baseline and one year after COVID-19

The CovILD cohort was recruited between April and June 2020. Out of 145 participants enrolled, 74 individuals with COVID-19-related persistent somatic symptoms, lung CT, LFT or cardiological abnormalities and complete BIPQ were included in the analysis. The major reasons of patient dropout were missing follow-up visits and incomplete BIPQ (**Figure 1**). The participants were predominantly male (65%), the median age at COVID-19 diagnosis was 56 years (IQR: 47 - 68), over one-third were active or ex-smokers (38%). Most participants suffered from comorbidities (74%), with cardiovascular disease, metabolic and respiratory disorders as the leading conditions. The participants were classified by COVID-19 severity as ambulatory (20%), moderate (hospitalized, no intensive care, no oxygen therapy, 54%) and severe COVID-19 survivors (hospitalized, oxygen therapy or intensive care, 26%). The ambulatory COVID-19 subset had the lowest median age, rate of smokers and comorbidity frequency (**Table 1**).

**Table 1:** Baseline demographic and clinical characteristic of the study cohort.

| Variable | Cohort | Ambulatory COVID-19 | Moderate COVID-19 | Severe COVID-19 | Significance<sup>a</sup> | Effect size<sup>a</sup> |
| --- | --- | --- | --- | --- | --- | --- |
| n, participants | 74 | 15 | 40 | 19 |  |  |
| Male sex | 65% (n = 48) | 33% (n = 5) | 70% (n = 28) | 79% (n = 15) | p = 0.013 | V = 0.34 |
| Age, years | median: 56<br>[IQR: 47 - 68]<br>range: 19 - 87 | median: 45<br>[IQR: 36 - 55]<br>range: 19 - 70 | median: 62<br>[IQR: 53 - 73]<br>range: 27 - 87 | median: 54<br>[IQR: 50 - 62]<br>range: 44 - 72 | p = 0.0013 | $\eta^2 = 0.16$ |
| Smoking history<sup>b</sup> | 38% (n = 28) | 13% (n = 2) | 48% (n = 19) | 37% (n = 7) | ns (p = 0.066) | V = 0.27 |
| Weight class<sup>c</sup> | normal: 38% (n = 28)<br>overweight: 43% (n = 32)<br>obesity: 19% (n = 14) | normal: 60% (n = 9)<br>overweight: 27% (n = 4)<br>obesity: 13% (n = 2) | normal: 30% (n = 12)<br>overweight: 50% (n = 20)<br>obesity: 20% (n = 8) | normal: 37% (n = 7)<br>overweight: 42% (n = 8)<br>obesity: 21% (n = 4) | ns (p = 0.37) | V = 0.17 |
| Comorbidity present | 74% (n = 55) | 47% (n = 7) | 80% (n = 32) | 84% (n = 16) | p = 0.022 | V = 0.32 |
| Cardiovascular disease | 43% (n = 32) | 13% (n = 2) | 48% (n = 19) | 58% (n = 11) | p = 0.024 | V = 0.32 |
| Hypertension | 30% (n = 22) | 13% (n = 2) | 30% (n = 12) | 42% (n = 8) | ns (p = 0.19) | V = 0.21 |
| Metabolic disease | 38% (n = 28) | 13% (n = 2) | 45% (n = 18) | 42% (n = 8) | ns (p = 0.089) | V = 0.26 |
| Hypercholesterolemia | 22% (n = 16) | 0% (n = 0) | 32% (n = 13) | 16% (n = 3) | p = 0.026 | V = 0.31 |
| Type II diabetes | 14% (n = 10) | 6.7% (n = 1) | 7.5% (n = 3) | 32% (n = 6) | p = 0.028 | V = 0.31 |
| Gastrointestinal disease | 14% (n = 10) | 0% (n = 0) | 20% (n = 8) | 11% (n = 2) | ns (p = 0.14) | V = 0.23 |
| Malignancy | 12% (n = 9) | 6.7% (n = 1) | 18% (n = 7) | 5.3% (n = 1) | ns (p = 0.31) | V = 0.18 |
| Respiratory disease | 24% (n = 18) | 13% (n = 2) | 28% (n = 11) | 26% (n = 5) | ns (p = 0.54) | V = 0.13 |
| Chronic kidney disease | 6.8% (n = 5) | 0% (n = 0) | 7.5% (n = 3) | 11% (n = 2) | ns (p = 0.46) | V = 0.14 |
| Immune deficiency | 4.1% (n = 3) | 0% (n = 0) | 2.5% (n = 1) | 11% (n = 2) | ns (p = 0.23) | V = 0.2 |
| WHO COVID-19 severity | median: 4<br>[IQR: 3 - 4.8]<br>range: 2 - 7 | median: 2<br>[IQR: 2 - 2]<br>range: 2 - 2 | median: 4<br>[IQR: 3 - 4]<br>range: 3 - 4 | median: 6<br>[IQR: 6 - 6]<br>range: 5 - 7 | p < 0.001 | $\eta^2 = 0.86$ |

<sup>a</sup>COVID-19 severity strata comparison; categorical variables: $\chi^2$ test with Cramer V effect size statistic, numeric variables: Kruskal-Wallis test with $\chi^2$ effect size statistic
<sup>b</sup>Former or active smoker
<sup>c</sup>Overweight: body mass index 25 - 30 kg/m<sup>2</sup>, obesity: > 30 kg/m<sup>2</sup>

Nearly three-quarter of participants (72%) suffered from persistent somatic symptoms at one year after COVID-19, with significant fatigue (41%), reduced physical performance (35%), sleep disorders (32%) and exertional dyspnea (22%) being most frequent. The symptom frequencies and fatigue rating were comparable between the COVID-19 severity strata (**Supplementary Table S2**). LFT abnormalities were discerned in 32% of participants and tended to be most common in severe COVID-19 survivors. Residual lung CT lesions were found in 54% individuals and their frequency and scoring was significantly higher in moderate and severe COVID-19 than in the ambulatory disease subset. The most common cardiological finding was low grade diastolic dysfunction (64% of cohort), which was significantly more frequent in moderate and severe than in ambulatory COVID-19 survivors. Nearly 80% of severe disease survivors attended COVID-19-specific rehabilitation, the rehabilitation rates in the remaining severity strata were below 20% (**Supplementary Table S2**). Most laboratory parameters at one year after COVID-19 were within their normal values. Mild anemia and improper glycemia control (HbA1c) were evident solely in moderate and severe COVID-19 survivors (**Supplementary Table S3**).

### Illness perception at one year after COVID-19

As measured by Cronbach’s alpha (42), the BIPQ tool had an acceptable internal consistency in the study cohort (*α* = 0.8, 95% CI: 0.69 - 0.86) (**Supplementary Figure S1**). The consequences, timeline, identity, concern and emotional representation items were strongly positively inter-correlated. Significant, moderate-to-strong positive association was observed between the personal control and treatment control components as well as between the coherence and treatment control items.

The median illness perception score defined as the sum of all items (15) was 23 (IQR: 15 - 32) in the study collective, the differences between ambulatory, moderate and severe COVID-19 were not significant. The treatment control and coherence BIPQ items were rated the highest followed by the personal control and emotional representation components. Significant differences between the COVID-19 severity strata were detected for the consequences, concern, emotional representation and coherence BIPQ items, which peaked in severe COVID-19 convalescents (**Table 2**).

**Table 2:**
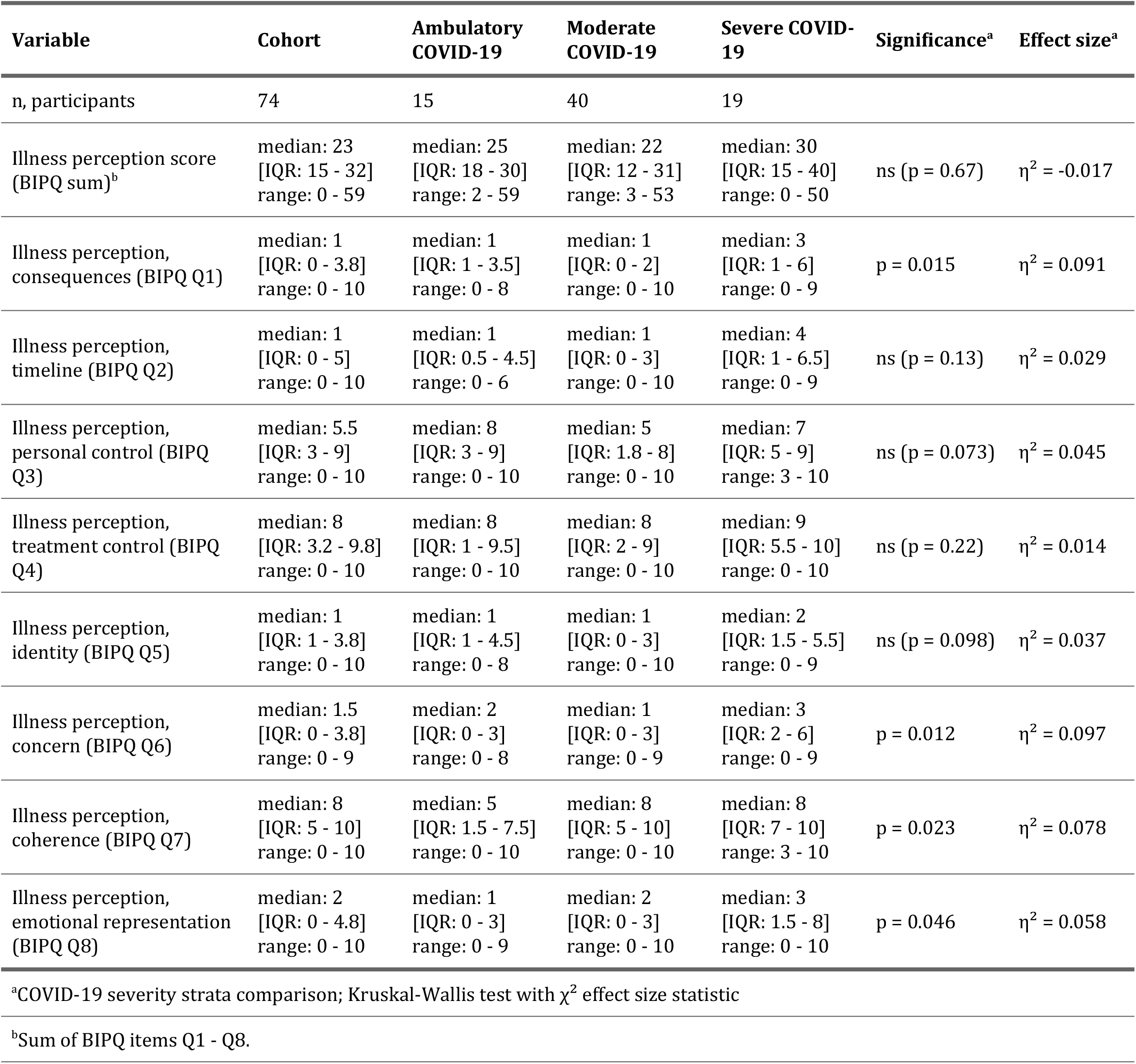
Illness perception score and item values of the Brief Illness Perception Questionnaire (BIPQ, Q1 - Q8) at one year after COVID-19.

### Key factors for overall illness perception

To identify the most important factors among 65 candidate explanatory variables (**Supplementary Table S1**) influencing the illness perception score at one year after COVID-19, three penalized multi-parameter regression algorithms: Elastic Net (34), LASSO (36) and Bayesian LASSO (37) were employed. The final models explained at least 47% and 36% of the illness perception score variance in the entire data set and 10-fold cross-validation, respectively (**Supplementary Figure S2**).

The number of variables with non-zero coefficients varied between 16 for the Elastic Net and 5 for the Bayesian LASSO regression (**Figure 2A**). The strongest positive correlates of the illness perception score in Elastic Net and LASSO modeling were pre-existing immune deficiency and respiratory disease as well as hair loss, fatigue rating and reduced physical performance at the one-year follow up. Fatigue scoring and respiratory comorbidity were the major positive correlates in the Bayesian LASSO model (**Supplementary Figures S2 - S5**). Of note, neither age, sex, acute COVID-19 severity nor LFT, CT and TTE abnormalities were selected as non-zero model coefficients by any of the regression algorithms. Their effects on the illness perception scoring was not significant in a direct analysis either (**Supplementary Figure S6**).

**Figure 2.**
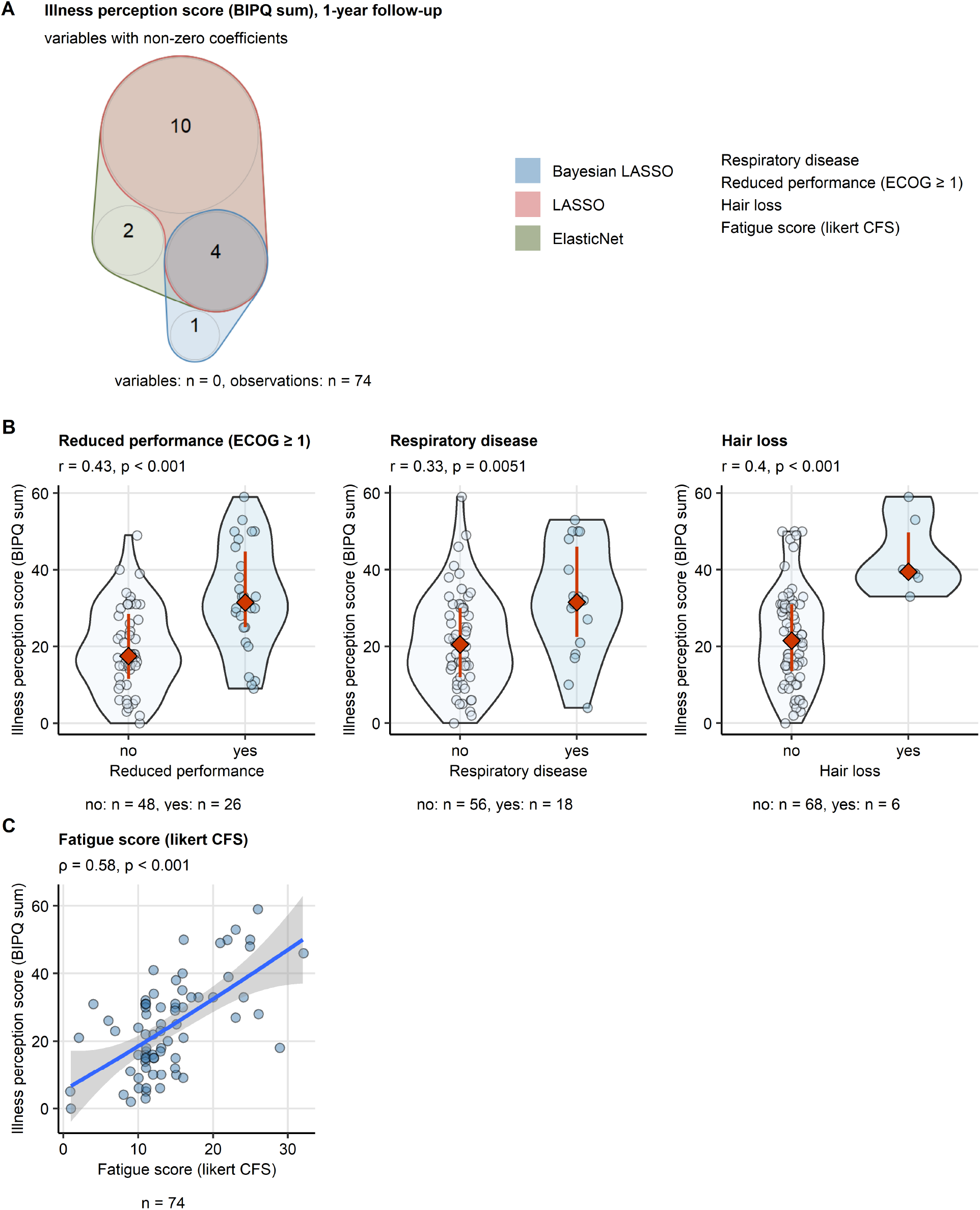
Key factors associated with disease perception one year after COVID-19. Illness perception score (sum of BIPQ items) at one year after COVID-19 was modeled as a function of 65 candidate independent variables using the Elastic Net, LASSO and Bayesian LASSO algorithms. Key factors affecting the illness perception score were identified as independent variables with non-zero coefficients in all three models. (A) Numbers of variables with non-zero coefficients identified by each algorithm presented in a quasi-proportional Venn diagram. The key factors are listed next to the diagram. (B) Relationship between the illness perception score and the categorical key factors: reduced physical performance (Eastern Cooperative Oncology Group [ECOG] score > 0) and hair loss at one year after COVID and respiratory comorbidity was investigated by Mann-Whitney test with Wilcoxon r effect size statistic. Effect size statistic and p values are indicated in the plot captions. Illness perception score values are presented in violin plots. Points represent single observations. Red diamonds and whiskers depict medians and interquartile ranges. Numbers of complete observations are displayed under the plots. (C) Relationship between the illness perception score and likert Chalder’s fatigue score (CFS) was investigated by Spearman’s correlation. The correlation coefficient (ρ) and p value are indicated in the plot caption. Each point represents a single observations. The blue line with gray ribbon depict the fitted second order terms and 95% confidence intervals. The number of complete observations is displayed under the plot.

The key variables associated with the illness perception rating selected by all three models were (1) reduced physical performance, (2) hair loss and (3) fatigue rating at the one-year follow-up as well as (4) baseline respiratory comorbidity (**Figure 2A**). Each of those parameters was found significantly associated with higher illness perception scores in a direct comparison or correlation analysis (**Figure 2BC**).

### Heterogeneity of illness perception

Three subsets of participants which differed qualitatively in BIPQ components, termed further ‘illness perception clusters’, were identified by PAM clustering (**Supplementary Figure S7**). Roughly half of study participants (cluster #1, 51% of participants) had low scoring of the emotional representation, concern, identity, timeline and consequences components along with high rating of the coherence, personal control and treatment control items. This translated to a low median illness perception score (15, IQR: 9 - 21). Another 27% of participants assigned to the cluster #2 were characterized by low levels of self-perceived personal or treatment control of their COVID-19 sequelae but otherwise by low rating of the concern, identity, timeline, consequences and emotional representation items. The cluster #3 individuals (22% of participants) had highly elevated rating of the concern, identity, timeline, consequences and emotional representation components as compared with the clusters #1 and #2. As a result, the cluster #3 displayed the highest median illness perception score (44, IQR: 37 - 50) (**Figure 3AB**).

**Figure 3.**
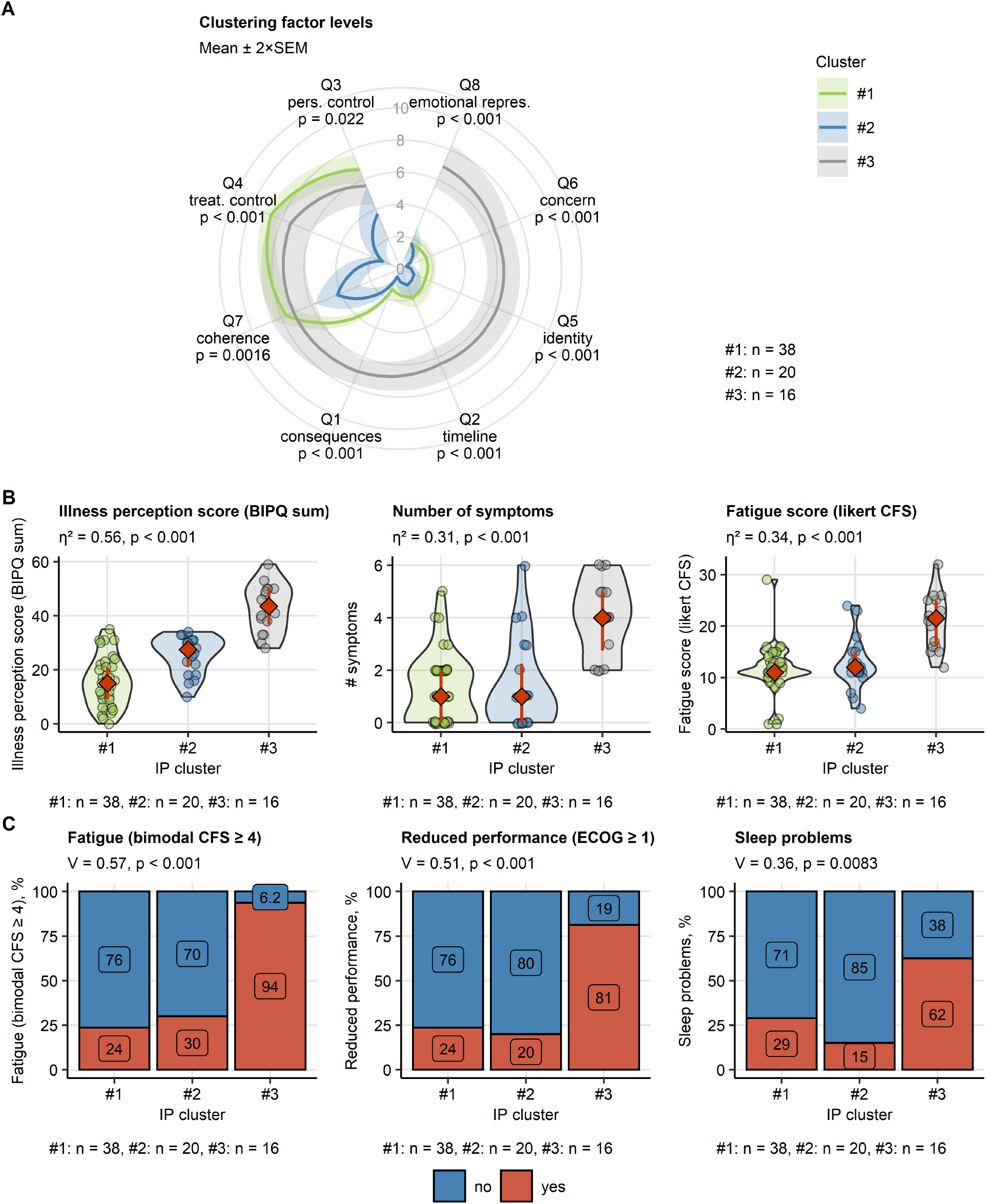
Heterogeneity of illness perception and residual symptoms one year after COVID-19. Three subsets of study participants (illness perception clusters) were identified by clustering in respect to the Brief Illness Perception Questionnaire items (BIPQ, Q1 - Q8) with the PAM (partitioning around medoids) clustering algorithm and Euclidean distance metric. Numbers of observations assigned to the clusters are displayed under the plots. (A) Mean BIPQ item scores at one year after COVID-19 in the illness perception clusters. Statistical significance was determined by Kruskal-Wallis test. P values are indicated below the variable names. Lines represent mean values, tinted ribbons depict 2×SEM (standard error of the mean) intervals. (B) Illness perception score (sum of BIPQ items), number of symptoms and likert Chalder’s fatigue score (CFS) at one year after COVID-19 in the illness perception clusters. Statistical significance was determined by Kruskal-Wallis test with η^2^ effect size statistic. Effect size statistic and p values are indicated in the plot captions. Response values are presented in violin plots. Points represent single observations. Red diamonds and whiskers depict medians and interquartile ranges. (C) Frequencies of significant fatigue (bimodal CFS ≥ 4), reduced physical performance (Eastern Cooperative Oncology Group [ECOG] score > 0) and sleep problems at one year after COVID-19 in the illness perception clusters. Statistical significance was determined by χ^2^ test with Cramer’s V effect size statistic. Effect size statistic and p values are indicated in the plot captions.

Among baseline demographic and clinical parameters and follow-up readouts of somatic complaints and cardiopulmonary abnormalities (**Supplementary Table S1**), COVID-19-related persistent somatic symptoms demonstrated the largest significant differences between the illness perception clusters. In particular, the cluster #3 individuals with high level of disease-related concerns suffered from multiple persistent somatic symptoms, persistent fatigue, physical performance loss, sleep problems, dyspnea and hair loss at one year after COVID-19. The cluster #3 comprised predominantly of individuals with higher COVID-19 severity (WHO grade), lung CT abnormalities and the peak rates of COVID-19-specific rehabilitation. The persistent somatic symptom frequency and intensity was comparable in the clusters #1 and #2. Yet, in the cluster #2 characterized by low self-perceived disease coherence and control, frequencies of smokers, metabolic comorbidity and LFT abnormalities at one year after COVID-19 tended to be higher than in the cluster #1. The rehabilitation rate in the cluster #2 was significantly lower than in the cluster #1 (**Figure 3** and **Supplementary Table S4**).

## Discussion

By penalized multi-parameter regression (34,36,37) applied to 65 candidate explanatory variables including demographic, clinical, somatic symptom data, laboratory and cardiopulmonary assessment results, we could discern 4 features explaining 47% of overall illness perception variance at one year after COVID-19. Those factors were: fatigue scoring, reduced physical performance and hair loss at the one-year follow-up and pre-existing respiratory comorbidity. Of note, the effects of age, sex, COVID-19 severity or residual lung lesions in CT, LFT findings and heath abnormalities in TTE on long-term illness perception were negligible. Furthermore, high levels of COVID-19-related concerns, consequences and emotional representation was observed primarily in the subset of participants with multiple residual somatic symptoms and moderate-to-severe acute COVID-19 course.

Literature on illness perception in COVID-19 patients is scarce (17,26–28,43). To our best knowledge, this is the first report assessing severity and components of illness perception in COVID-19 convalescents with persistent somatic symptoms or cardiopulmonary findings at the clinical assessment one year after diagnosis. The link between disease severity, symptoms, therapy control and illness perception is well founded in other chronic conditions (12,15,44). In particular, we identified fatigue as a strong covariate of overall illness perception scoring after COVID-19 and as an important factor characterizing individuals with high levels of COVID-19-related concerns, emotional representation and consequences in clustering analysis. Similar effects of chronic fatigue were described in arthritis (12) and hematological malignancy (44). Both acute COVID-19 and its post-acute sequelae encompass various respiratory symptoms such as cough or dyspnea (1,2,4,7). In addition, COVID-19 was found to exacerbate symptoms and worsen disease control in asthma (45). Such superimposed COVID-19-dependent and independent airway manifestations may hence explain more severe illness perception in respiratory comorbidity in our study cohort.

By clustering analysis we could identify three subsets of participants differing in key illness perception components. Half of individuals displayed low severity of overall illness perception, good self-perceived coherence, personal and treatment control paralleled by low burden of persistent somatic symptoms. Another 27% of participants showed a similarly low level of persistent somatic symptoms or fatigue. Yet their illness perception was hallmarked by poor disease understanding and disbelief in personal and treatment control. By contrast, the remaining minor cluster suffered from multiple somatic complaints at the one-year follow-up and was enriched in individuals with residual lung lesions in CT, severe COVID-19 course, significant fatigue, sleep problems and hair loss.

Their illness perception was characterized primarily by intense emotional representation, concern, burden of consequences and disease identity. This latter subset is of particular interest and concern for psychological and psychiatric management of post COVID-19 syndrome as high scoring of emotional components of illness perception was correlated with signs of shame, guilt, stress, depression and anxiety both in the general population during the pandemic (25) and in acute COVID-19 (26–28). The individuals in this cluster could potentially profit most from psychological and psychiatric interventions.

Collectively, we demonstrate an important effect of persistent somatic symptoms rather than clinical assessment results revealing lung lesions, lung function deficits or heart abnormalities on severity and quality of illness perception at one year after COVID-19. In the long run, this aspect may bear consequences for the patient’s physical and mental health following COVID-19 and public health in general. Negative illness perceptions were found to accompany somatic symptom disorder, perpetuate symptoms in somatoform disorders and to predict higher future healthcare expenditure (46). In multiple aspects, post COVID-19 condition resembles persistent somatic manifestations of functional or somatic symptom disorders (47).

Our study bears limitations. The most important one was the low participant number and substantial participant dropout due to missing follow-up visits and the BIPQ answers, which may have resulted in a selection bias. Furthermore, longitudinal rating of illness perception or inclusion of a general population control (22,23,25,48) would allow us to assess possible improvement or worsening and explore factors associated with illness perception at consecutive time points. The study collective was recruited during the pandemic onset before introduction of effective anti-viral drugs and vaccination. We were hence unable to assess the effect of improved treatment and prevention on illness perception (24). Finally, the study variable set lacks parameters which may be vital for severity and character of illness perception such as family status, education and COVID-19 knowledge (17), media consumption (49) and quarantine duration (28).

## Conclusion

Our results demonstrate, that persistent somatic symptoms rather than clinical assessment results, revealing lung lesions, lung function deficits or heart abnormalities, impact on severity and quality of illness perception at one year after COVID-19. Hence, it is to account not only for acute COVID-19 severity but also for the interplay between persistent somatic symptoms and individual illness perceptions when allocating rehabilitation and psychological or psychiatric resources.

## Supporting information

STROBE checklist

COI forms

## Data Availability

An R data (RDa) file with anonymized patient data will be made available upon request to the corresponding author. The study pipeline is available at https://github.com/PiotrTymoszuk/CovILD-IPQ.

## Acknowledgments

We acknowledge commitment of the study participants and the medical staff during the COVID-19 pandemic.

