## Supplementary Material for "Persistent somatic symptoms are key to individual illness perception at one year after COVID-19"

CovILD study team

2022-09-05

### Supplementary Methods

#### Study cohort and approval

The CovILD longitudinal observation cohort study (ClinicalTrials.gov: NCT04416100) includes adult (≥ 18 year) patients with a symptomatic, PCR-confirmed SARS-CoV-2 infection. The follow-up visits were scheduled at two, three, six months and one year after COVID-19 diagnosis. A total of 190 individuals were screened for participation. In the current report, a subset of the original CovILD cohort was investigated, which displayed (1) persistent COVID-19-related symptoms or (2) any abnormality in chest computed tomography (CT) or (4) any deficits in lung function testing (LFT) or (5) any cardiological abnormality in trans-thoracic echocardiography (TTE) at the one-year follow-up. The final analysis inclusion criterion was availability of complete Brief Illness Perception Questionnaire (IPQ) data (1).

n = 145 participants were recruited between March and June 2020 at three centers located in Tyrol, Austria: the Medical University of Innsbruck (Innsbruck), St. Vinzenz Hospital (Zams) and Karl-Landsteiner Rehabilitation Facility (Muenster). The reasons for screening failure were denied informed consent and self-declared logistic and temporary incompatibility with the scheduled study visits. Out of n = 74 participants, who completed the one-year follow-up, n = 74 suffered from symptoms or displayed cardiopulmonary abnormalities (CT, LFT or TTE findings), filled out the BIPQ and had a complete set of modeling variables (**Supplementary Table S1**). The recruitment process scheme is presented in **Figure 1**. Baseline characteristic of the study cohort is presented in **Table 1**, follow-up features are summarized in **Supplementary Tables S2** (symptoms, cardiopulmonary findings) and **S3** (laboratory parameters). BIPQ item scores and the sum BIPQ score are summarized in **Table 2**.

The study was performed in accordance with the Declaration of Helsinki and the European Data Policy. All participants gave written informed consent. The study protocol was approved by the ethics committee at the Medical University of Innsbruck (approval number 1103/2020).

#### Procedures and variables

The protocol of each follow-up visit included a survey of COVID-19-related symptoms, blood laboratory parameter assessment, LFT, TTE and chest CT as described in more detail before (2–4). Comorbidities and data on acute COVID-19 course (symptoms, laboratory parameters, functional tests and imaging) were recorded retrospectively at the two-month follow-up visit based on information provided by the participant and electronic patient records.

#### Baseline demographic and clinical status, acute COVID-19 severity

The baseline demographic variables included sex (male/female), age at COVID-19 diagnosis, smoking history, body weight and height. The following pre-existing comorbidities were surveyed: obesity, metabolic disease, cardiovascular disease, hypertension, type II diabetes, hypercholesterolemia, gastrointestinal disease, malignancy, chronic kidney disease, immune deficiency, pulmonary and respiratory conditions. Among the surveyed comorbidities, asthma, COPD (chronic obstructive pulmonary disease), interstitial lung disease and other chronic pulmonary disorders were subsumed under the 'respiratory disease' variable included in the independent modeling variable set. Study participants were stratified according to acute COVID-19 severity as ambulatory (outpatient, WHO grade 1 - 2), moderate (hospitalized at normal infection ward, no oxygen therapy, WHO grade 3 - 4) and severe COVID-19 survivors (hospitalized with oxygen therapy or mechanical ventilation or intensive care, WHO grade 5 - 7).

#### Cardiopulonary assessment

LFT abnormality was defined as FVC (forced vital capacity) < 80% or FEV1 (forced expiratory volume in 1 second) < 80% or TLC (total lung capacity) < 80% or DLCO (diffusion lung capacity for carbon monoxide) < 80% of the predicted reference value or FEV1:FVC ratio < 70% of the predicted reference value (3). CT images were screened for ground glass opacities (GGO), consolidations, bronchiectasis and reticulations according to the Fleischner Society glossary (5). Chest CT abnormalities were scored separately for each lobe with the CT severity score (2–4): 0 - no abnormality, 1 - minimal (subtle GGO), 2 - mild (several GGO, subtle reticulation), 3 - moderate (multiple GGO, reticulation, small consolidation), 4 - severe (extensive GGO, consolidation, reticulation with distortion), 5 - massive (multiple findings, parenchymal destruction). The sum for all five lobes was used in the analysis. Any chest CT abnormality is defined as CT severity score ≥ 1. The most frequent cardiological abnormality in TTE was low grade diastolic dysfunction (**Supplementary Table S2**). No reduced left ventricular ejection fraction was observed in the participants included in the current analysis.

#### COVID-19-related persistent somatic symptoms and exertional capacity

The following features were surveyed as COVID-19-related persistent somatic symptoms:

- **reduced physical performance** assessed with the ECOG scale (Eastern Cooperative Oncology Group). Reduced physical performance was defined as ECOG ≥ 1

- **dyspnea** rated with the mMRC scale (Modified Medical British Research Council). Dyspnea was defined as mMRC ≥ 1
- **cough**, self-reported, surveyed as a yes/no item
- **sleep problems**, self-reported, surveyed as a yes/no item
- **night sweating**, self-reported, surveyed as a yes/no item
- **hyposmia or anosmia**, self-reported, surveyed as a yes/no item
- **dermatological symptoms**, self-reported, surveyed as a yes/no item
- **gastrointestinal symptoms**, self-reported, surveyed as a yes/no item
- **hair loss**, self-reported, surveyed as a yes/no item
- **significant fatigue**, measured with the bimodal Chalder's Fatigue Scale (CFS). Significant fatigue was defined as bimodal CFS ≥ 4 (6,7)

The individual counts of persistent somatic symptoms were subsequently calculated. Since only few participants displayed values of ECOG ≥ 2 or mMRC ≥ 2, the dichotomous 'reduced performance (ECOG ≥ 1)' and 'dyspnea (mMRC ≥ 1)' independent variables were used in modeling instead of the full numeric scales. Fatigue intensity was rated with the likert Chalder's Fatigue Scale (CFS) (6,7). Exertional capacity was assessed by six-minute walking distance (SMWD) test performed according to the ATS guidelines (8). The reference SMWD values were obtained as described by Crapo and colleagues (8) and differences between the actual and reference values were calculated.

#### Laboratory parameters

Laboratory parameters encompassing hemoglobin (total and glycated hemoglobin [HbA1c]), biomarkers of iron turnover (ferritin [FT], transferrin saturation [TF-Sat], hepcidin and soluble transferrin receptor [sTFR]), inflammation and coagulation (C-reactive protein [CRP] and D-dimer) and cardiovascular pathology (N-terminal pro-brain natriuretic peptide [NT-proBNP]) were determined routinely at the certified laboratory of the Central Institute for Medical and Chemical Diagnostic at the Federal University Hospital in Innsbruck. Other markers of systemic inflammation, interleukin 6 (IL6) and procalcitonin, were within the normal ranges in the analysis collective.

#### Illness perception

Illness perception was scored with the Brief Illness Perception Questionnaire (IPQ) developed by Broadbent and colleagues (1). Each of the 8 BIPQ items (questions Q1 - Q8) was scored in with an 11-point likert scale:

- **Q1 consequences** (0: no consequences, 10: very severe consequences)
- **Q2 timeline** (0: very short, 10: forever)
- **Q3 personal control** (0: no control, 10: extreme control)
- **Q4 treatment control** (0: no control, 10: extreme control)

- **Q5 identity** (0: no complaints, 10: multiple severe complaints)
- **Q6 concerns** (0: no concerns at all, 10: extreme concerns)
- **Q7 coherence** (0: not at all, 10: I understand my illness very good)
- **Q8 emotional representation** (0: no emotions at all, 10: extreme emotions)

The illness perception score was calculated as an arithmetic sum of items Q1 - Q8, with the negative items Q3, Q4 and Q7 inverted. The BIPQ tool demonstrated acceptable consistency as measured by Cronbach's $\alpha$ = 0.8 (bootstrap percentile 95% CI: [0.69 - 0.86]) (**Supplementary Figure S1**) (9).

Variables used in the exploratory data analysis and multi-parameter modeling are listed in **Supplementary Table S1**.

#### Data import, transformation and visualization methods

Data import, transformation, statistical analysis and result visualization was done with R version 4.2.0 (R Foundation for Statistical Computing). For import of data tables and variable transformation, the *tidyverse* package bundle (10) and *rlang* package (11) was used. The *ggplot2* (12) and *cowplot* (13) packages and the development package *figur* (https://github.com/PiotrTymoszuk/figur) were used for data visualization. Quasi-proportional Venn diagrams were generated with the *nVennR* package (14). The manuscript and supplementary material files were created with the *rmarkdown* (15), *knitr* (16), *bookdown* (17) packages, *Affiliations* Pandoc filter set (https://github.com/drdanholmes/Affiliations) and the development package *figur* (https://github.com/PiotrTymoszuk/figur).

#### Hypothesis testing

For statistical hypothesis testing, base R functions, the package *rstatix* (18) and development package *exda* (https://github.com/PiotrTymoszuk/ExDA) were used. Statistical significance of differences in frequency of categorical variable levels was assessed by $\chi^2$ test with Cramer V effect size statistic. Since multiple study variables were non-normally distributed as investigated by Shapiro-Wilk test and quantile-quantile plots, differences in numeric variables were assessed by non-parametric Mann-Whitney (effect size: Wilcoxon r) or Kruskal-Wallis test (effect size: $\eta^2$), as appropriate. Correlations were analyzed with Spearman test (function *cor.test()*, package *stats*) and visualized by fitting a second order trend (function *geom_smooth()*, package *ggplot*, formula: $y \sim x + I\left(x^2\right)$, development package *exda*).

#### BIPQ tool consistency

Spearman's pair-wise correlations between the BIPQ items were calculated (function *cor.test()*, package *stats*) and visualized in a bubble plot (**Supplementary Figure S1**). Cronbach's $\alpha$ was calculated with package *psych* (9,19). 95% confidence intervals for $\alpha$ were obtained by the bootstrap technique (n = 1000).

#### Multi-parameter modeling

In multi-parameter modeling, a set of 65 independent variables was used. The response, illness perception score defined as a sum of all BIPQ items, was square-root transformed to guarantee normal distribution. For numeric variables, both first- and second-order terms were included to account for non-linear relationship between the independent variable and response. During pre-processing, numeric variables (independent and responses) were normalized with mean centering (Z-scores). The full independent variable list is provides in **Supplementary Table S1**.

Three L1 penalized regression algorithms were utilized to model illness perception score at one year after COVID-19: Elastic Net (20), LASSO (least absolute shrinkage and selection operator) (21) and Bayesian LASSO (22). The choice of penalized regression over canonical backward elimination or forward selection was motivated by highly multi-dimensional nature of the data set.

The Elastic Net and LASSO algorithms are implemented in R by the *glmnet* package (23). The optimal $\lambda$ parameters for Elastic Net and LASSO regression were found as values associated with the minimum model deviance in 200-repeats 10-fold cross-validation (CV, function *cv.glmnet()*, Elastic Net: $\alpha$ = 0.5, LASSO: $\alpha$ = 1, $\lambda$ parameter 'lambda.min' was extracted). The optimal $\lambda$ values were 0.15 for the Elastic Net and 0.076 for the LASSO model. Finally, Elastic Net and LASSO models with the optimal $\lambda$ values were constructed with *caret* package (function *train()*) (24) and their performance in the training data set and 10-repeats 10-fold CV was tested (**Supplementary Figure D2**). Estimates of non-zero final model coefficients (**Supplementary Figures S3** and **S4**) were extracted with the *coef* method (s parameter set to the optimal $\lambda$ value).

The Bayesian LASSO algorithm is implemented in R by the *monomvn* package (25). The Bayesian Lasso model was constructed with the *caret* package (function *train()*, method: 'blasso', iteration number: T = 1000). The optimal value of the sparsity parameter, which controls the fraction of non-zero coefficients in the final model based on the posterior distributions, was found by 10-repeats 10-fold CV with the RMSE (root mean squared error) statistic as a selection criterion (**Supplementary Figure S2**). The optimal sparsity value selected from the 0.1 - 0.7 range was 0.4. The model coefficient matrix X was extracted from the 'finalModel' slot of the caret model. The coefficient values

(**Supplementary Figure S5**) were calculated as matrix column medians over all algorithm iterations (22).

Extraction of caret model performance measures: RMSE and $R^2$ defined as $1 - MSE/Var(y)$ (**Supplementary Figure S2**) was accomplished by the in-house-developed package *caretExtra* (https://github.com/PiotrTymoszuk/caretExtra). Model assumption testing (normality of residuals, Shapiro-Wilk test, method *residuals()*) and visual quality control based on plots of residuals versus fitted and residuals quantile-quantile plots was done with the *caretExtra* package (method *plot()*).

The key factors impacting on the illness perception score at one year after COVID-19 (respiratory disease, reduced performance (ECOG ≥ 1), hair loss and fatigue score (likert CFS)) were defined as variables with non-zero coefficients in all multi-parameter models.

#### Clustering analysis

Study participants were clustered in respect to BIPQ items (consequences (BIPQ Q1), timeline (BIPQ Q2), personal control (BIPQ Q3), treatment control (BIPQ Q4), identity (BIPQ Q5), concern (BIPQ Q6), coherence (BIPQ Q7) and emotional representation (BIPQ Q8)) by the PAM algorithm (partition around medoids, package *cluster*) (26) with the Euclidean distance measure between the observations (package *philentropy*) (27). The BIPQ item scores were not pre-processed, since they were measured with the same likert scale. The PAM/Euclidean distance procedure was chosen based on the high explained clustering variance (ratio of the between-cluster sum of squares to the total sum of squares) and stability (cluster assignment accuracy in 10-fold CV, cluster assignment by 7-nearest neighbors label propagation algorithm) (28,29) as compared with several other clustering algorithms (**Supplementary Figure S7A**). The choice of cluster number (k = 3) was based on the bend of the curve of within-cluster sum of squares (generated with package *factoextra*, **Supplementary Figure S7B**) (30) and visual analysis of the distance heat map (**Supplementary Figure S7C**). Clustering object development, clustering variance calculation, cross-validation and visualization were performed with the in-house developed package *clustTools* (https://github.com/PiotrTymoszuk/clustTools).

#### Data and code availability

An R data (RDa) file with anonymized patient data will be made available upon request to the corresponding author. The study pipeline is available at https://github.com/PiotrTymoszuk/CovILD-IPQ.

#### Supplementary Tables

***Supplementary Table S1: Study variables***

| R name | Plot/table label | Unit | Description |
| --- | --- | --- | --- |
| sex | Sex | | |
| age | Age | years | Age during COVID-19 |
| age_sq | $Age^2$ | years | Age during COVID-19, second order |
| WHO | WHO COVID-19 severity | | WHO severity grade, acute COVID-19 |
| WHO_sq | WHO COVID-19 $severity^2$ | | WHO severity grade, acute COVID-19, second order |
| cat_WHO | COVID-19 severity | | A: ambulatory, HM: hospitalized, moderate, HS: hospitalized, severe |
| rehabilitation | Rehabilitation | % | COVID-19 specific rehabilitation |
| weight_class | Weight class | | Normal: BMI ≤ 25 $kg/m^2$, overweight: 25 – 30 $kg/m^2$, obesity: > 30 $kg/m^2$ |
| smoking_history | Smoking history | % | Smoking history |
| lufo_red | LFT abnormality | % | Lung function testing abnormality: FVC < 80% or FEV1 < 80% or TLC < 80% or DLCO < 80% predicted or FEV1:FVC < 70% predicted reference value |
| Hb | Hb | g/dL | Blood hemoglobin |
| Hb_sq | $Hb^2$ | g/dL | Blood hemoglobin, second order |
| anemia | Anemia | % | Females: Hb < 120 g/dL, males: Hb < 140 g/dL |
| ferritin | FT | µg/L | Serum ferritin |
| ferritin_sq | $FT^2$ | µg/L | Serum ferritin, second order |
| TSAT | TF-Sat | % | Transferrin saturation |
| TSAT_sq | $TF\text{-}Sat^2$ | % | Transferrin saturation, second order |
| sTFR | sTFR | mg/L | Serum soluble transferrin receptor |
| sTFR_sq | $sTFR^2$ | mg/L | Serum soluble transferrin receptor, second order |
| Hepcidin | Hepcidin | ng/mL | Serum hepcidin |

| R name | Plot/table label | Unit | Description |
|---|---|---|---|
| Hepcidin_sq | $Hepcidin^2$ | ng/mL | Serum hepcidin, second order |
| NTproBNP | NT-proBNP | pg/mL | Serum N-terminal pro-brain natriuretic peptide |
| DDimer | D-dimer | µg/L | Serum D-dimer |
| DDimer_sq | $D\text{-}dimer^2$ | µg/L | Serum D-dimer, second order |
| CRP | CRP | mg/L | Serum C-reactive protein |
| CRP_sq | $CRP^2$ | mg/L | Serum C-reactive protein, second order |
| HbA1c | HbA1c | % | Blood glycated hamoglobin |
| HbA1c_sq | $HbA1c^2$ | % | Blood glycated hamoglobin, second order |
| diastolic_dysf | Diastolic dysfunction | % | Heart diastolic dysfunction, trans-thoracic echocardiography |
| no_comorb | Number of comorbidities | | Number of comorbidities |
| no_comorb_sq | $(\text{Number comorbidities})^2$ | | Number of comorbidities, second order |
| comorb_present | Comorbidity present | % | Comorbidity present |
| cardiovascular_comorb | Cardiovascular disease | % | Cardiovascular disease |
| hypertension_comorb | Hypertension | % | Hypertension |
| respi_comorb | Respiratory disease | % | Respiratory disease: asthma, chronic obstructive pulmonary disease, interstitial lung disease or other lung disorder |
| endometabolic_comorb | Metabolic disease | % | Metabolic disease |
| hyperchol_comorb | Hypercholesterolemia | % | Hypercholesterolemia |
| diabetes_comorb | Type II diabetes | % | Type II diabetes |
| ckd_comorb | Chronic kidney disease | % | Chronic kidney disease |
| gastro_comorb | Gastrointestinal disease | % | Gastrointestinal disease |
| malingancy_comorb | Malignancy | % | Malignancy |

| R name | Plot/table label | Unit | Description |
|---|---|---|---|
| immdef_comorb | Immune deficiency | % | Immune deficiency |
| sympt_present | Symptoms present | % | Symptoms present at the follow-up visit |
| sympt_number | Number of symptoms | | Number of symptoms at the follow-up visit |
| sympt_number_sq | (Number of symptoms)$^2$ | | Number of symptoms at the follow-up visit, second order |
| sleep_sympt | Sleep problems | % | Self-reported sleep problems at the follow-up visit (yes/no) |
| dyspnoe_sympt | Dyspnea (mMRC ≥ 1) | % | Dyspnea at the follow-up visit (Modified Medical Research Council score > 0) |
| cough_sympt | Cough | % | Self-reported cough at the follow-up visit (yes/no) |
| night_sweat_sympt | Night sweat | % | Self-reported night sweating at the follow-up visit (yes/no) |
| gastro_sympt | Gastrointestinal symptoms | % | Self-reported gastrointestinal symptoms at the follow-up visit (yes/no) |
| anosmia_sympt | Hypo/anosmia | % | Self-reported hyposmia or anosmia at the follow-up visit (yes/no) |
| fatigue_sympt | Reduced performance (ECOG ≥ 1) | % | Reduced physical performance at the follow-up visit (Eastern Cooperative Oncology Group > 0) |
| ct_severity_score | CT severity score | | Chest computer tomography severity score |
| ctss_sq | (CT severity score)$^2$ | | Chest computer tomography severity score, second order |
| ct_severity_any | CT abnormality (CT score ≥ 1) | % | Any abnormality in chest CT, CT severity score ≥ 1 |
| hair_loss_sympt | Hair loss | % | Hair loss at the follow-up visit |
| derma_sympt | Dermatological symptoms | % | Dermatological symptoms at the follow-up visit |

| R name | Plot/table label | Unit | Description |
|---|---|---|---|
| smwd | SMWD | m | Six-minute walking distance |
| smwd_sq | $SMWD^2$ | m | Six-minute walking distance, second order |
| smwd_dref | SMWD vs reference | m | Six-minute walking distance, difference of the actual and reference value |
| smwd_dref_sq | $(SMWD\ vs\ reference)^2$ | m | Six-minute walking distance, difference of the actual and reference value, second order |
| Chalder_FS | Fatigue score (likert CFS) | | Fatigue score, likert Chalder's Fatigue Score |
| Chalder_FS_sq | $(Fatigue\ score\ (likert\ CFS))^2$ | | Fatigue score, likert Chalder's Fatigue Score, second order |
| Chalder_FS_bimodal | Fatigue (bimodal CFS ≥ 4) | % | Significant fatigue, bimodal Chalder's Fatigue Score ≥ 4 |
| ipq_total | Illness perception score (BIPQ sum) | | Illness perception score, Illness Perception Questionnaire, item sum |
| ipq_q1 | Illness perception, consequences (BIPQ Q1) | | Illness perception, consequences item, Illness Perception Questionnaire Question 1 |
| ipq_q2 | Illness perception, timeline (BIPQ Q2) | | Illness perception, timeline item, Illness Perception Questionnaire Question 2 |
| ipq_q3 | Illness perception, personal control (BIPQ Q3) | | Illness perception, personal control item, Illness Perception Questionnaire Question 3 |
| ipq_q4 | Illness perception, treatment control (BIPQ Q4) | | Illness perception, treatment control item, Illness Perception Questionnaire Question 4 |
| ipq_q5 | Illness perception, identity (BIPQ Q5) | | Illness perception, identity item, Illness Perception Questionnaire Question 5 |
| ipq_q6 | Illness perception, concern (BIPQ Q6) | | Illness perception, concern item, Illness Perception Questionnaire Question 6 |

| R name | Plot/table label | Unit | Description |
|---|---|---|---|
| ipq_q7 | Illness perception, coherence (BIPQ Q7) | | Illness perception, coherence, Illness Perception Questionnaire Question 7 |
| ipq_q8 | Illness perception, emotional representation (BIPQ Q8) | | Illness perception, emotional representation, Illness Perception Questionnaire Question 8 |

***Supplementary Table S2: COVID-19 symptoms, performance, fatigue, exertional capacity, cardiopulmonary abnormalities and illness perception at the one-year follow-up.***

| Variable | Cohort | Ambulatory COVID-19 | Moderate COVID-19 | Severe COVID-19 | Significance[a] | Effect size[a] |
|---|---|---|---|---|---|---|
| n, participants | 74 | 15 | 40 | 19 | | |
| Symptoms present | 72% (n = 53) | 80% (n = 12) | 72% (n = 29) | 63% (n = 12) | ns (p = 0.55) | V = 0.13 |
| Number of symptoms | median: 2 [IQR: 0 - 3] range: 0 - 6 | median: 2 [IQR: 1 - 4] range: 0 - 6 | median: 1.5 [IQR: 0 - 3] range: 0 - 6 | median: 2 [IQR: 0 - 3] range: 0 - 6 | ns (p = 0.7) | $\eta^2$ = -0.018 |
| Reduced performance (ECOG ≥ 1)[b] | 35% (n = 26) | 33% (n = 5) | 35% (n = 14) | 37% (n = 7) | ns (p = 0.98) | V = 0.025 |
| Dyspnea (mMRC ≥ 1)[c] | 22% (n = 16) | 13% (n = 2) | 22% (n = 9) | 26% (n = 5) | ns (p = 0.65) | V = 0.11 |
| Cough | 12% (n = 9) | 6.7% (n = 1) | 15% (n = 6) | 11% (n = 2) | ns (p = 0.68) | V = 0.1 |
| Sleep problems | 32% (n = 24) | 47% (n = 7) | 25% (n = 10) | 37% (n = 7) | ns (p = 0.28) | V = 0.19 |
| Night sweat | 18% (n = 13) | 33% (n = 5) | 15% (n = 6) | 11% (n = 2) | ns (p = 0.18) | V = 0.21 |
| Hypo/anosmia | 14% (n = 10) | 20% (n = 3) | 18% (n = 7) | 0% (n = 0) | ns (p = 0.13) | V = 0.23 |
| Dermatological symptoms | 9.5% (n = 7) | 6.7% (n = 1) | 7.5% (n = 3) | 16% (n = 3) | ns (p = 0.55) | V = 0.13 |
| Gastrointestinal symptoms | 2.7% (n = 2) | 6.7% (n = 1) | 2.5% (n = 1) | 0% (n = 0) | ns (p = 0.49) | V = 0.14 |
| Hair loss | 8.1% (n = 6) | 13% (n = 2) | 7.5% (n = 3) | 5.3% (n = 1) | ns (p = 0.68) | V = 0.1 |
| Fatigue score (likert CFS)[d] | median: 12 [IQR: 11 - 16] range: 1 - 32 | median: 14 [IQR: 11 - 19] range: 2 - 26 | median: 12 [IQR: 11 - 15] range: 1 - 24 | median: 13 [IQR: 11 - 19] range: 1 - 32 | ns (p = 0.54) | $\eta^2$ = -0.011 |
| Fatigue (bimodal CFS ≥ 4)[d] | 41% (n = 30) | 53% (n = 8) | 32% (n = 13) | 47% (n = 9) | ns (p = 0.29) | V = 0.18 |
| LFT abnormality[e] | 32% (n = 24) | 20% (n = 3) | 30% (n = 12) | 47% (n = 9) | ns (p = 0.21) | V = 0.2 |
| CT abnormality (CT score ≥ 1)[f] | 54% (n = 40) | 13% (n = 2) | 52% (n = 21) | 89% (n = 17) | p < 0.001 | V = 0.52 |
| CT severity score[g] | median: 1 [IQR: 0 - 5] range: 0 - 14 | median: 0 [IQR: 0 - 0] range: 0 - 5 | median: 1 [IQR: 0 - 2.2] range: 0 - 8 | median: 5 [IQR: 2.5 - 10] range: 0 - 14 | p < 0.001 | $\eta^2$ = 0.35 |
| Diastolic dysfunction | 64% (n = 47) | 27% (n = 4) | 68% (n = 27) | 84% (n = 16) | p = 0.0019 | V = 0.41 |
| SMWD, m[h] | median: 560 [IQR: 500 - 630] range: 270 - 760 | median: 620 [IQR: 560 - 650] range: 400 - 740 | median: 550 [IQR: 480 - 630] range: 270 - 760 | median: 560 [IQR: 500 - 640] range: 410 - 700 | ns (p = 0.11) | $\eta^2$ = 0.034 |

| **Variable** | **Cohort** | **Ambulatory COVID-19** | **Moderate COVID-19** | **Severe COVID-19** | **Significance[a]** | **Effect size[a]** |
|---|---|---|---|---|---|---|
| SMWD vs reference, m[i] | median: 0 [IQR: -61 - 46] range: -230 - 140 | median: 0 [IQR: -55 - 31] range: -230 - 120 | median: 0 [IQR: -56 - 46] range: -220 - 130 | median: 0 [IQR: -71 - 42] range: -210 - 140 | ns (p = 0.85) | $\eta^2 = -0.024$ |
| Rehabilitation | 31% (n = 23) | 6.7% (n = 1) | 18% (n = 7) | 79% (n = 15) | p < 0.001 | V = 0.61 |

[a]COVID-19 severity strata comparison; categorical variables: $\chi^2$ test with Cramer V effect size statistic, numeric variables: Kruskal-Wallis test with $\chi^2$ effect size statistic

[b]Eastern Cooperative Oncology Group performance score

[c]Modified Medical Research Council dyspnea score

[d]Chalder's Fatigure Score

[e]Lung function testing abnormality: FVC < 80% or FEV1 < 80% or TLC < 80% or DLCO < 80% predicted or FEV1:FVC < 70% predicted reference value

[f]Any abnormality in chest computed tomography (CT), CT severity score ≥ 1

[g]Chest computer tomography severity score

[h]Six minute walking distance

[i]Six-minute walking distance, difference between the actual and reference value

***Supplementary Table S3: Laboratory parameters at the one-year follow-up.***

| Variable | Cohort | Ambulatory COVID-19 | Moderate COVID-19 | Severe COVID-19 | Significance[a] | Effect size[a] |
|---|---|---|---|---|---|---|
| n, participants | 74 | 15 | 40 | 19 | | |
| Anemia[b] | 11% (n = 8) | 0% (n = 0) | 15% (n = 6) | 11% (n = 2) | ns (p = 0.28) | V = 0.19 |
| Hb, g/dL[c] | median: 150<br>[IQR: 140 - 160]<br>range: 110 - 180 | median: 140<br>[IQR: 140 - 150]<br>range: 130 - 160 | median: 150<br>[IQR: 140 - 160]<br>range: 110 - 170 | median: 160<br>[IQR: 150 - 160]<br>range: 140 - 180 | p = 0.0029 | $\eta^2$ = 0.14 |
| FT, µg/L[d] | median: 160<br>[IQR: 110 - 260]<br>range: 13 - 1300 | median: 140<br>[IQR: 48 - 170]<br>range: 13 - 450 | median: 170<br>[IQR: 130 - 270]<br>range: 13 - 1300 | median: 180<br>[IQR: 110 - 250]<br>range: 25 - 870 | ns (p = 0.19) | $\eta^2$ = 0.018 |
| TF-Sat, %[e] | median: 26<br>[IQR: 20 - 33]<br>range: 6 - 61 | median: 28<br>[IQR: 18 - 32]<br>range: 14 - 49 | median: 26<br>[IQR: 21 - 33]<br>range: 6 - 61 | median: 24<br>[IQR: 20 - 32]<br>range: 10 - 44 | ns (p = 0.82) | $\eta^2$ = -0.022 |
| sTFR, mg/L[f] | median: 3<br>[IQR: 2.4 - 3.4]<br>range: 1.6 - 6.2 | median: 2.8<br>[IQR: 2.3 - 3.2]<br>range: 2 - 4.1 | median: 3<br>[IQR: 2.4 - 3.3]<br>range: 1.6 - 6 | median: 3<br>[IQR: 2.5 - 3.4]<br>range: 1.9 - 6.2 | ns (p = 0.49) | $\eta^2$ = -0.008 |
| Hepcidin, ng/mL | median: 10<br>[IQR: 6.5 - 17]<br>range: 0 - 51 | median: 9.8<br>[IQR: 3 - 12]<br>range: 0 - 18 | median: 12<br>[IQR: 7 - 21]<br>range: 0 - 38 | median: 8.7<br>[IQR: 4.5 - 18]<br>range: 0 - 51 | ns (p = 0.28) | $\eta^2$ = 0.0078 |
| NT-proBNP, pg/mL[g] | median: 58<br>[IQR: 0 - 110]<br>range: 0 - 1600 | median: 0<br>[IQR: 0 - 73]<br>range: 0 - 140 | median: 68<br>[IQR: 0 - 150]<br>range: 0 - 1600 | median: 0<br>[IQR: 0 - 90]<br>range: 0 - 850 | ns (p = 0.18) | $\eta^2$ = 0.021 |
| D-dimer, µg/L | median: 320<br>[IQR: 210 - 530]<br>range: 0 - 4000 | median: 520<br>[IQR: 250 - 970]<br>range: 0 - 4000 | median: 320<br>[IQR: 210 - 590]<br>range: 170 - 1800 | median: 290<br>[IQR: 210 - 350]<br>range: 0 - 1100 | ns (p = 0.13) | $\eta^2$ = 0.029 |
| CRP, mg/L[h] | median: 0.14<br>[IQR: 0.07 - 0.3]<br>range: 0 - 7.4 | median: 0.11<br>[IQR: 0.06 - 0.28]<br>range: 0 - 0.53 | median: 0.14<br>[IQR: 0.07 - 0.3]<br>range: 0 - 2.4 | median: 0.16<br>[IQR: 0.035 - 0.31]<br>range: 0 - 7.4 | ns (p = 0.8) | $\eta^2$ = -0.022 |
| HbA1c, %[i] | median: 5.7<br>[IQR: 5.5 - 6]<br>range: 4.9 - 8.8 | median: 5.5<br>[IQR: 5.4 - 5.8]<br>range: 4.9 - 7.5 | median: 5.8<br>[IQR: 5.6 - 6.1]<br>range: 4.9 - 7.8 | median: 5.7<br>[IQR: 5.3 - 5.9]<br>range: 5 - 8.8 | ns (p = 0.11) | $\eta^2$ = 0.035 |

[a]COVID-19 severity strata comparison; categorical variables: $\chi^2$ test with Cramer V effect size statistic, numeric variables: Kruskal-Wallis test with $\chi^2$ effect size statistic

[b]Females: hemoglobin (Hb) < 120 g/dL, males: Hb < 140 g/dL

[c]Blood hemoglobin

[d]Serum ferritin

| Variable | Cohort | Ambulatory COVID-19 | Moderate COVID-19 | Severe COVID-19 | Significance[a] | Effect size[a] |
|---|---|---|---|---|---|---|
| [e]Serum transferrin saturation | | | | | | |
| [f]Serum soluble transferrin receptor | | | | | | |
| [g]Serum N-terminal pro-brain natriuretic peptide | | | | | | |
| [h]Serum C-reactive protein | | | | | | |
| [i]Blood glycated hamoglobin | | | | | | |

***Supplementary Table S4: Significant ($p < 0.05$) and near-significant ($p < 0.1$) differences between the illness perception clusters.***

| Variable | Cluster #1 | Cluster #2 | Cluster #3 | Significance[a] | Effect size[a] |
|---|---|---|---|---|---|
| n, participants | 38 | 20 | 16 | | |
| Smoking history | 26% (n = 10) | 55% (n = 11) | 44% (n = 7) | ns (p = 0.087) | V = 0.26 |
| Metabolic disease | 26% (n = 10) | 45% (n = 9) | 56% (n = 9) | ns (p = 0.087) | V = 0.26 |
| Respiratory disease | 16% (n = 6) | 25% (n = 5) | 44% (n = 7) | ns (p = 0.091) | V = 0.25 |
| WHO COVID-19 severity | median: 4 [IQR: 3 - 5] range: 2 - 7 | median: 3 [IQR: 2.8 - 4] range: 2 - 6 | median: 4 [IQR: 3 - 6] range: 2 - 7 | p = 0.048 | $\eta^2 = 0.058$ |
| Symptoms present | 63% (n = 24) | 65% (n = 13) | 100% (n = 16) | p = 0.017 | V = 0.33 |
| Number of symptoms | median: 1 [IQR: 0 - 2] range: 0 - 5 | median: 1 [IQR: 0 - 2.2] range: 0 - 6 | median: 4 [IQR: 2.8 - 5] range: 2 - 6 | p < 0.001 | $\eta^2 = 0.31$ |
| Reduced performance (ECOG ≥ 1)[b] | 24% (n = 9) | 20% (n = 4) | 81% (n = 13) | p < 0.001 | V = 0.51 |
| Dyspnea (mMRC ≥ 1)[c] | 11% (n = 4) | 20% (n = 4) | 50% (n = 8) | p = 0.0055 | V = 0.37 |
| Sleep problems | 29% (n = 11) | 15% (n = 3) | 62% (n = 10) | p = 0.0083 | V = 0.36 |
| Gastrointestinal symptoms | 0% (n = 0) | 0% (n = 0) | 12% (n = 2) | p = 0.024 | V = 0.32 |
| Hair loss | 0% (n = 0) | 0% (n = 0) | 38% (n = 6) | p < 0.001 | V = 0.57 |
| Fatigue score (likert CFS)[d] | median: 11 [IQR: 11 - 13] range: 1 - 29 | median: 12 [IQR: 11 - 15] range: 4 - 24 | median: 22 [IQR: 16 - 25] range: 12 - 32 | p < 0.001 | $\eta^2 = 0.34$ |
| Fatigue (bimodal CFS ≥ 4)[d] | 24% (n = 9) | 30% (n = 6) | 94% (n = 15) | p < 0.001 | V = 0.57 |
| LFT abnormality[e] | 21% (n = 8) | 45% (n = 9) | 44% (n = 7) | ns (p = 0.099) | V = 0.25 |
| CT severity score[f] | median: 1 [IQR: 0 - 5] range: 0 - 14 | median: 0 [IQR: 0 - 1.2] range: 0 - 5 | median: 4.5 [IQR: 0 - 10] range: 0 - 13 | p = 0.015 | $\eta^2 = 0.09$ |
| Rehabilitation | 32% (n = 12) | 10% (n = 2) | 56% (n = 9) | p = 0.012 | V = 0.35 |

[a]COVID-19 severity strata comparison; categorical variables: $\chi^2$ test with Cramer V effect size statistic, numeric variables: Kruskal-Wallis test with $\chi^2$ effect size statistic

[b]Eastern Cooperative Oncology Group performance score

[c]Modified Medical Research Council dyspnea score

| Variable | Cluster #1 | Cluster #2 | Cluster #3 | Significance[a] | Effect size[a] |
|---|---|---|---|---|---|
| [d]Chalder's Fatigure Score | | | | | |
| [e]Lung function testing abnormality: FVC < 80% or FEV1 < 80% or TLC < 80% or DLCO < 80% predicted or FEV1:FVC < 70% predicted reference value | | | | | |
| [f]Any abnormality in chest computed tomography (CT), CT severity score ≥ 1 | | | | | |

### Supplementary Figures

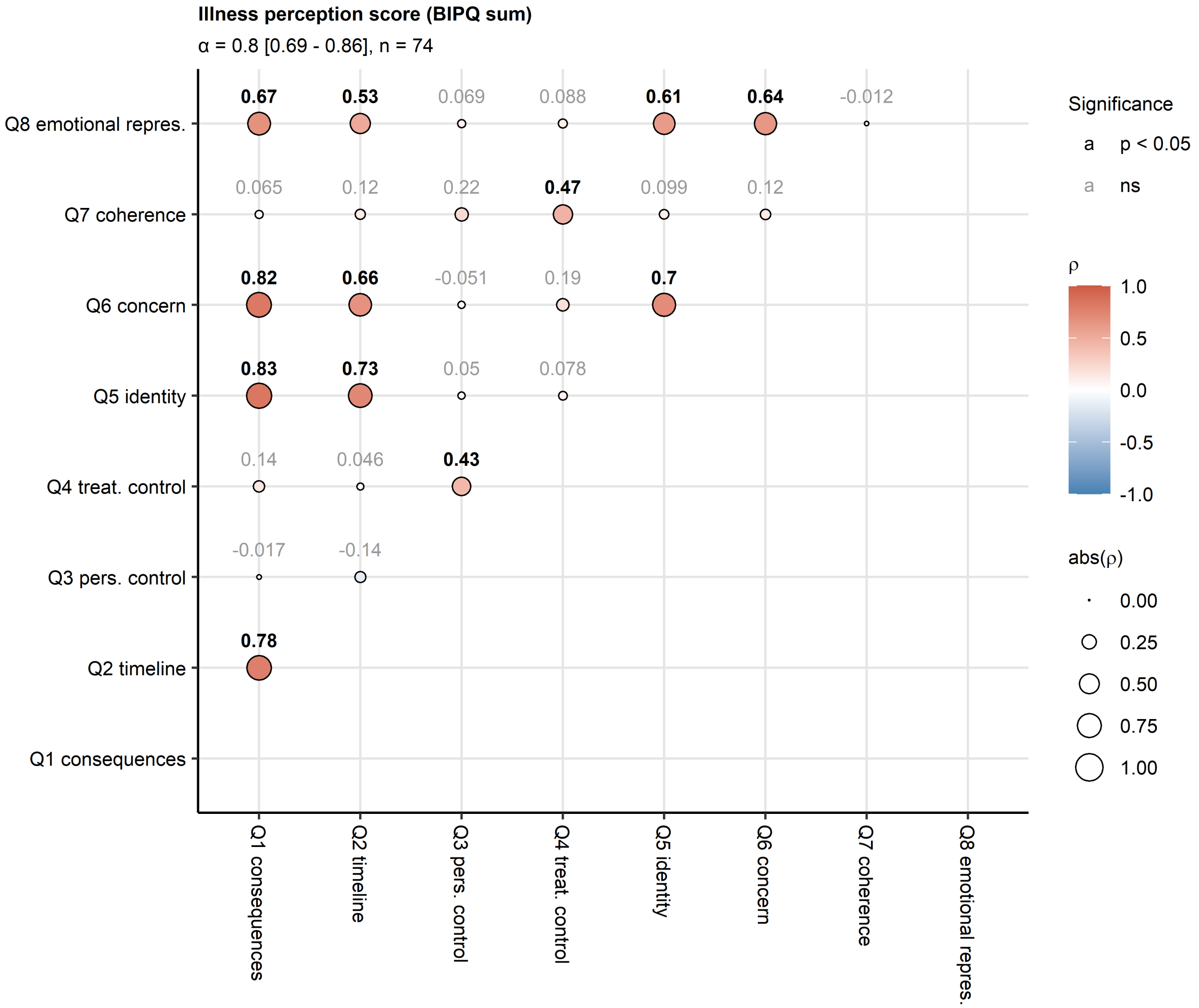


**Supplementary Figure S1. Coherence of the illness perception score.**

*Pair-wise correlation between particular items of the Brief Illness Perception Questionnaire (IPQ) was assessed by Spearman test and the results presented in the point plot. Point size corresponds to absolute values of the correlation coefficient ρ. Point color codes for the ρ value. The points are labeled with ρ values, significant effects are highlighted in bold. Internal consistency of the BIPQ tool was measured by Cronbach's α. The α statistic with 95% confidence intervals and the number of complete observations are displayed in the plot caption.*

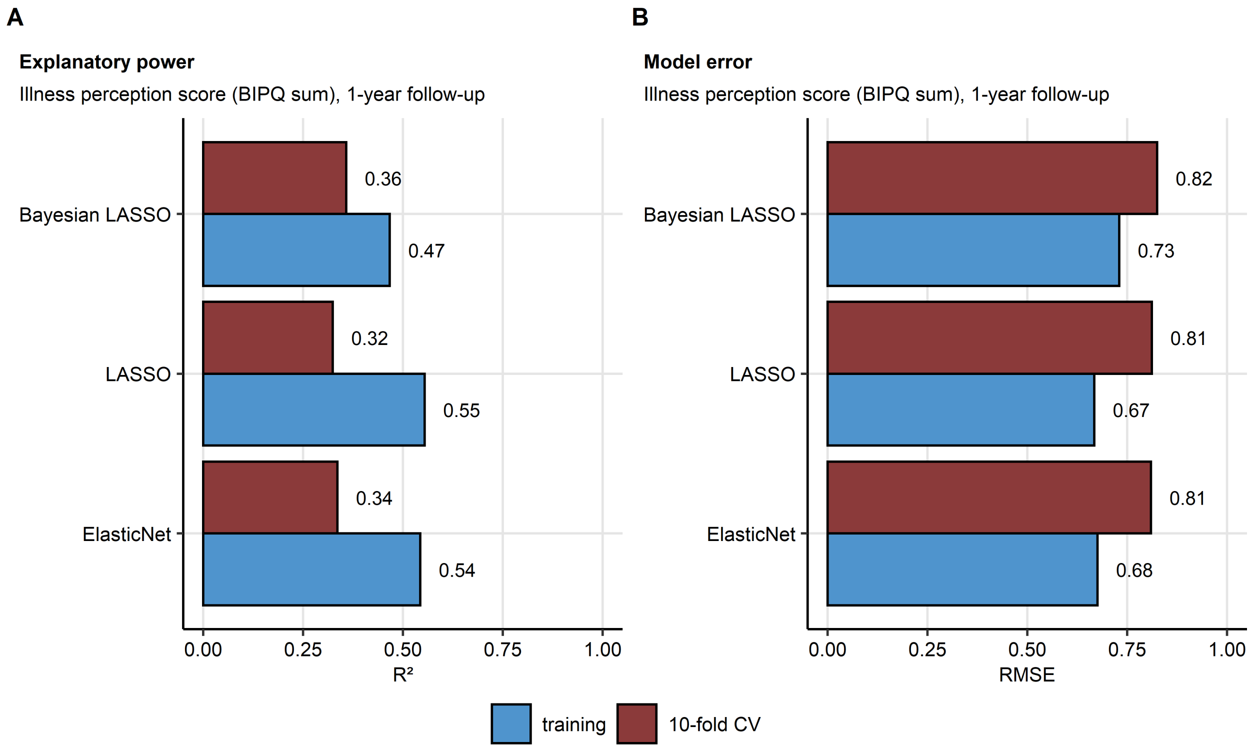


**Supplementary Figure S2. Multi-parameter model performance.**

*(A) Fraction of illness perception score variance explained by the Elastic Net, LASSO and Bayesian LASSO model in the training data set and 10-repeats 10-fold cross-validation (CV) was estimated by the* $R^2$ *statistic.*

*(B) Model error expressed as root mean squared error (RMSE) in the training and CV data sets.*

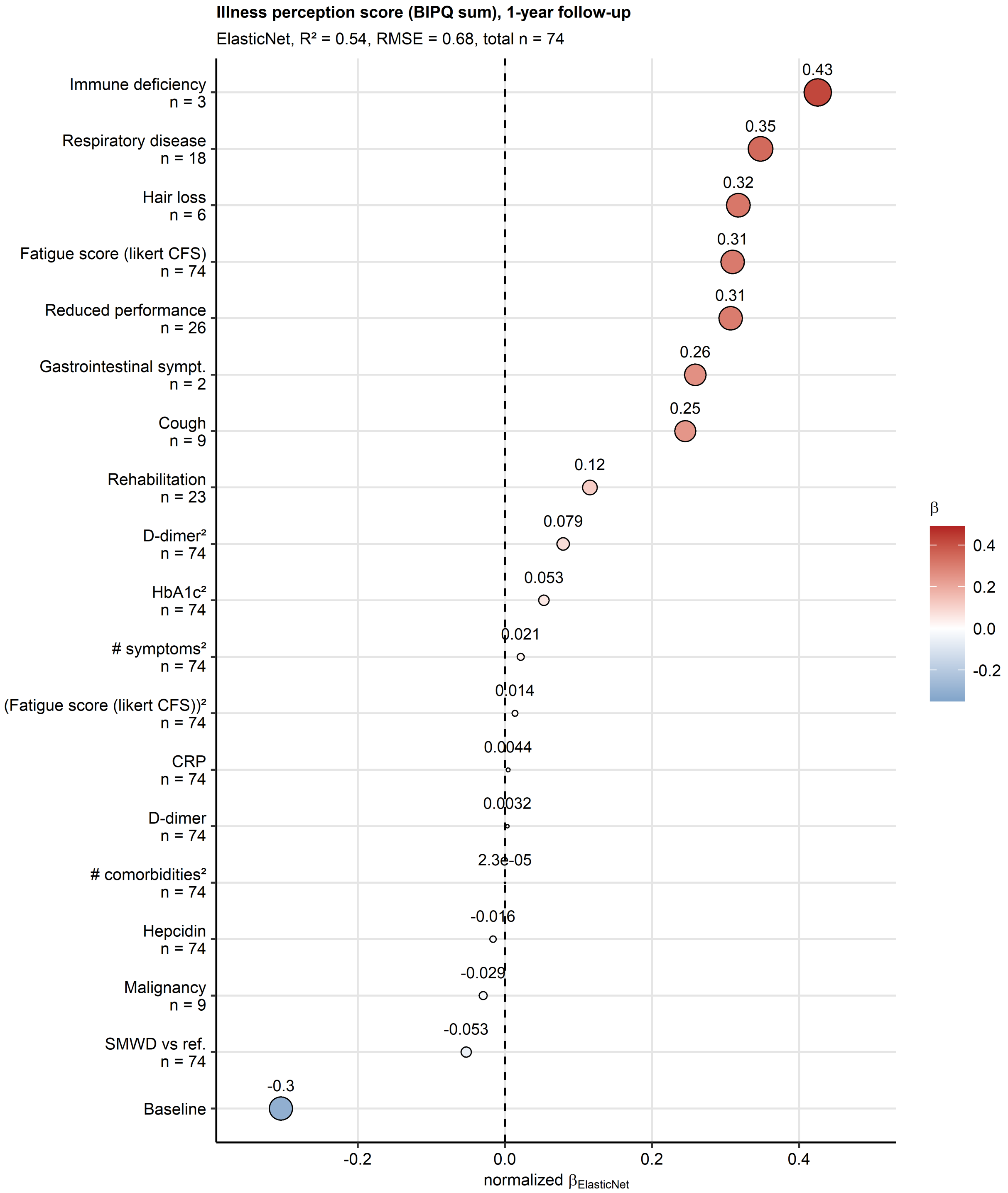


**Supplementary Figure S3. Non-zero coefficient estimates of the Elastic Net multi-parameter model.**

*Z-score normalized estimates (β) of the non-zero coefficients of the Elastic Net model are presented in the plot as points. Point color codes β value, point size corresponds to the absolute value of β. Model's performance measures in the training data set ($R^2$ and root mean squared error [RMSE]) and numbers of complete observations are indicated in the plot caption. The data points are labeled with their β values.*

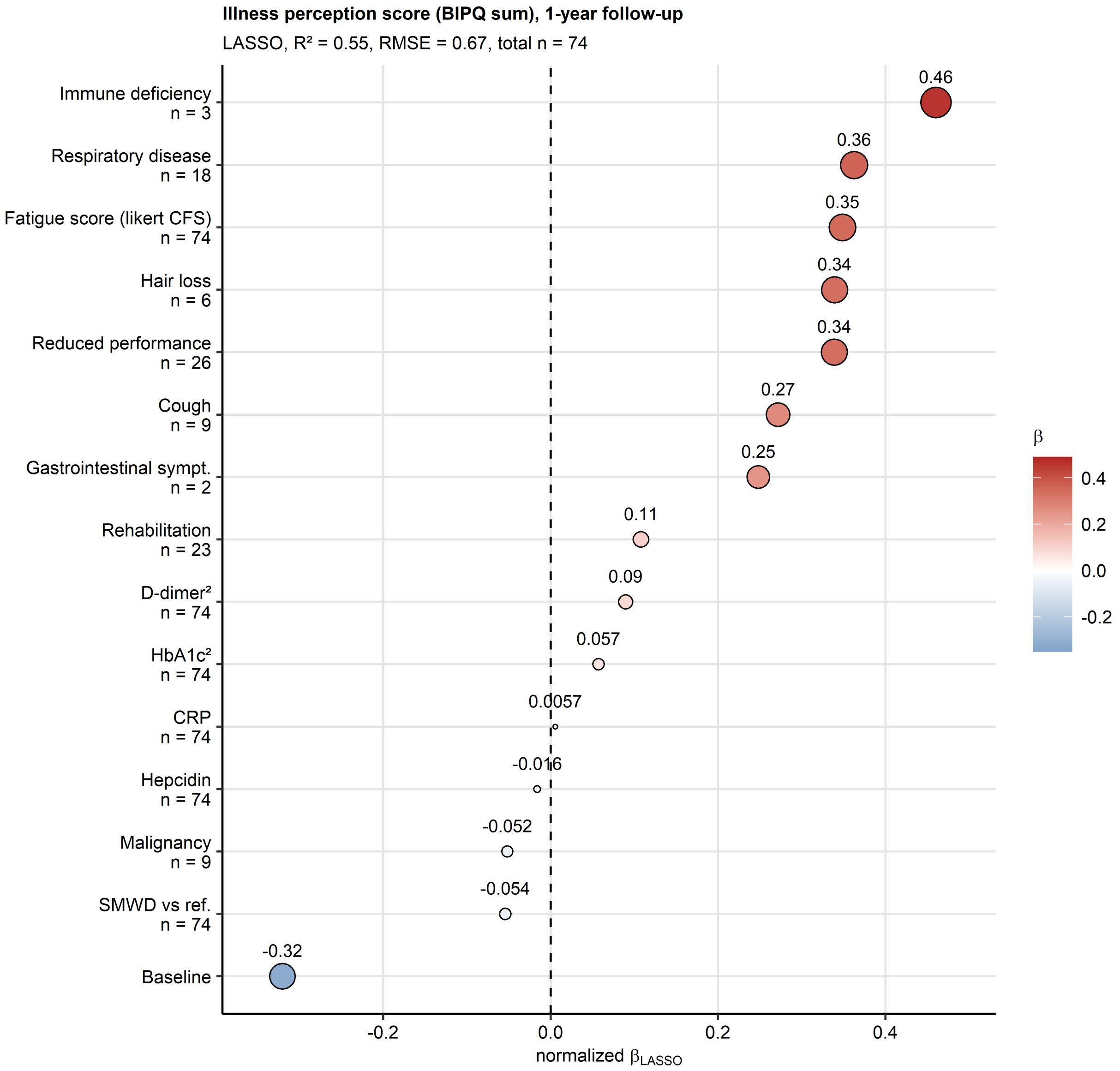


**Supplementary Figure S4. Non-zero coefficient estimates of the LASSO multi-parameter model.**

*Z-score normalized estimates (β) of the non-zero coefficients of the LASSO (least absolute shrinkage and selection operator) model are presented in the plot as points. Point color codes β value, point size corresponds to the absolute value of β. Model's performance measures in the training data set ($R^2$ and root mean squared error [RMSE]) and numbers of complete observations are indicated in the plot caption. The data points are labeled with their β values.*

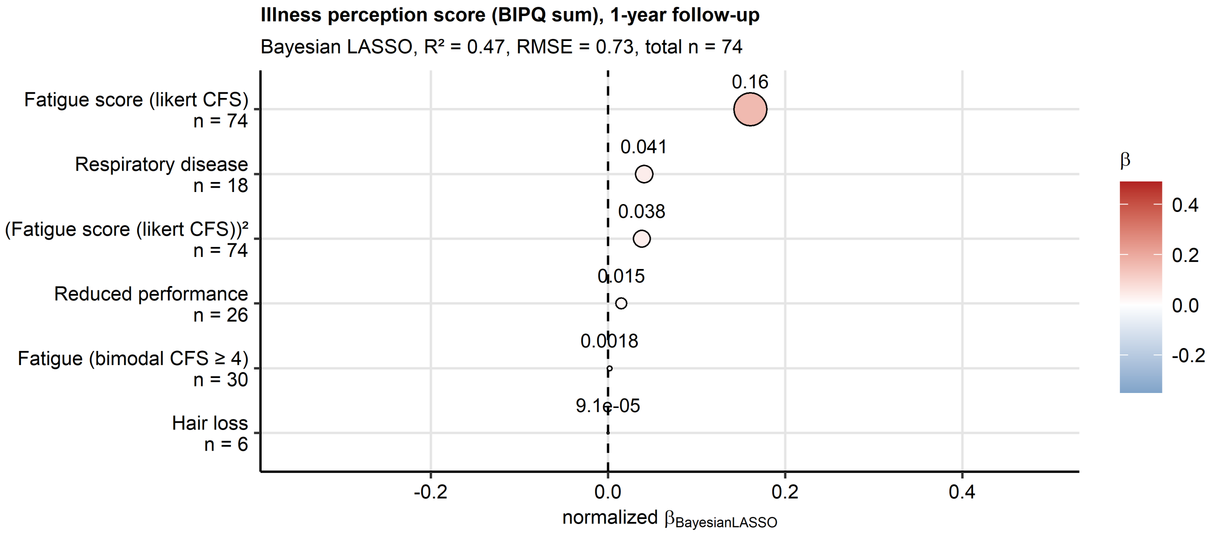


**Supplementary Figure S5. Non-zero coefficient estimates of the Bayesian LASSO multi-parameter model.**

*Z-score normalized estimates (β) of the non-zero coefficients of the Bayesian LASSO model are presented in the plot as points. Point color codes β value, point size corresponds to the absolute value of β. Model's performance measures in the training data set ($R^2$ and root mean squared error [RMSE]) and numbers of complete observations are indicated in the plot caption. The data points are labeled with their β values.*

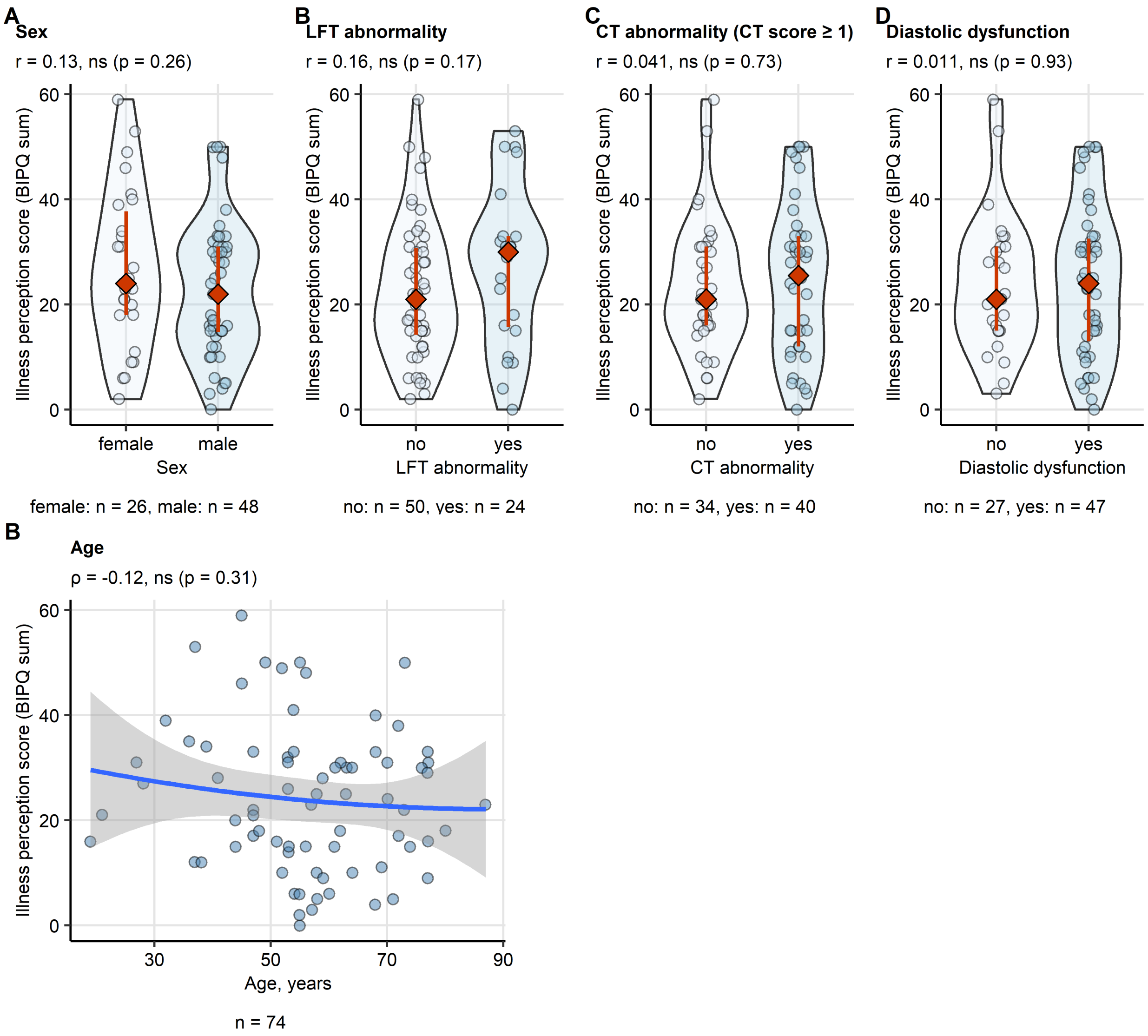


**Supplementary Figure S6. Age, sex and cardiopulmonary abnormalities at the one-year follow-up and illness perception..**

*(A - D) Differences in the illness perception score between males and female (A), the study participants with and without lung function (LFT) testing abnormality (B), chest computed tomography (CT) abnormality (C) and heart diastolic dysfunction (D) were assessed by Mann-Whitney test with Wilcoxon r effect size statistic. Effect size statistic and p values are indicated in the plot captions. Illness perception score values are presented i violin plots. Each point represents a single observation. Red diamonds and whiskers represent medians with interquartile ranges. Numbers of individuals with and without cardiopulmonary abnormalities are displayed under the plots.*

*(F). Correlation of age at COVID-19 diagnosis and the illness perception score at one year after COVID-19 diagnosis was investigated by Spearman's test. Correlation coefficients (ρ) and p values are indicated in the plot captions. Each point represents a single observations. Blue lines with gray ribbons depict the fitted second order terms and 95% confidence intervals. Numbers of complete observations are displayed under the plots.*

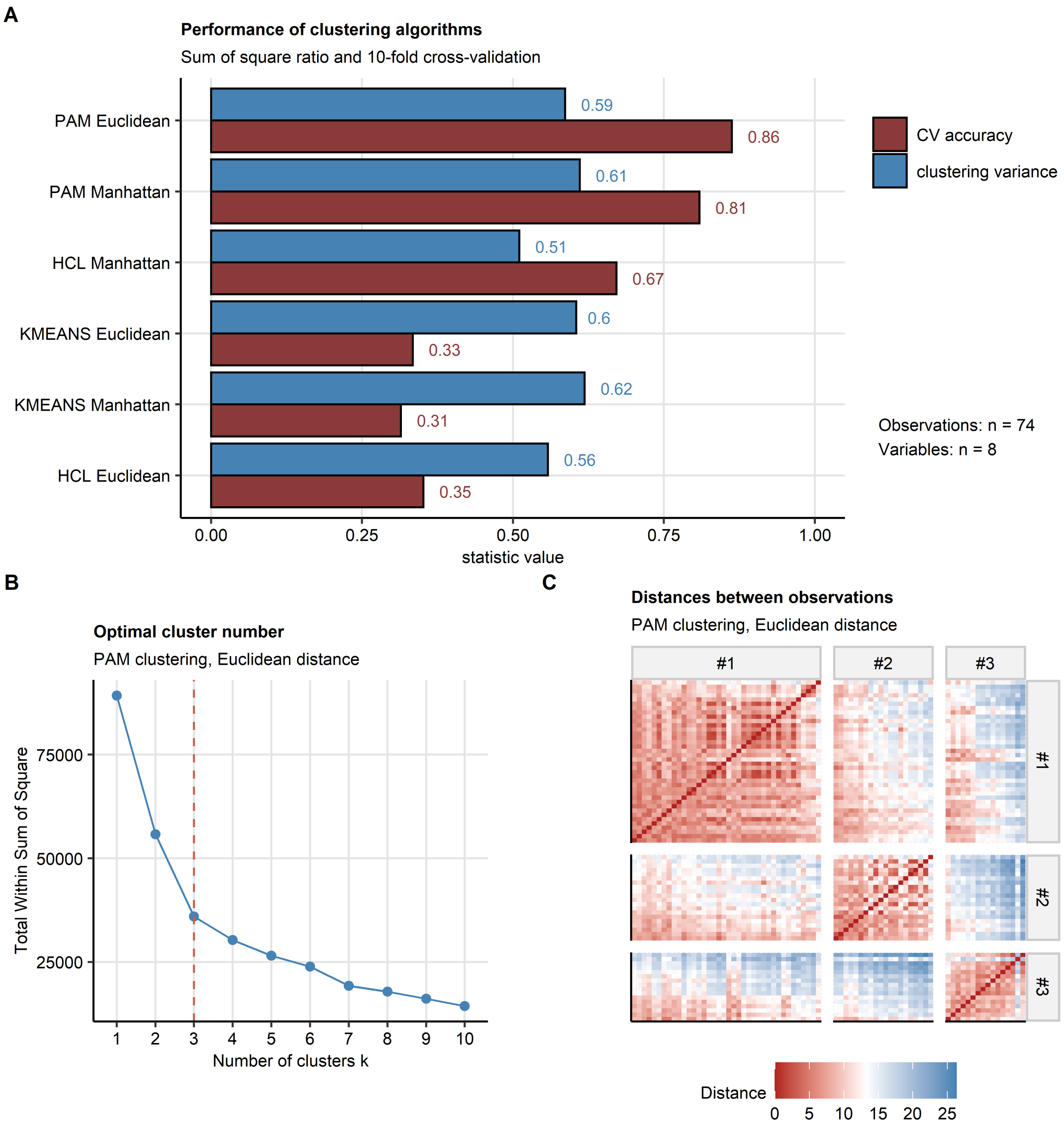


**Supplementary Figure S7. Choice of the optimal clustering algorithm and the cluster number.**

*Study participants were clustered in respect to the Brief Illness Perception Questionnaire items (Q1 - Q8) using the PAM (partitioning around medoids) clustering algorithm and Euclidean distance between the observations. Numbers of complete observations and clustering variables are indicated in (A)*

*(A) Comparison of clustering algorithm performance (PAM: partitioning around medoids, HCL: Ward D2 hierarchical clustering, KMEANS). Explained clustering variance was defined as a ratio of the between-cluster sum of squares to the total sum of squares, cluster assignment accuracy was investigated in 10-fold cross-validation (CV). Note the superior CV accuracy of the PAM/Euclidean distance.*

*(B, C) Choice of the optimal cluster number for the hierarchical clustering/Euclidean distance algorithm. The optimal cluster number was selected based on the bend of the curve of within-cluster sum of squares (B, dashed red line: the selected cluster number) and visual inspection of the distance heat map (C).*
